# Age-related heterogeneity in immune responses to SARS-CoV-2 vaccine BNT162b2

**DOI:** 10.1101/2021.02.03.21251054

**Authors:** Dami A. Collier, Isabella A.T.M. Ferreira, Prasanti Kotagiri, Rawlings Datir, Eleanor Lim, Emma Touzier, Bo Meng, Adam Abdullahi, The CITIID-NIHR BioResource COVID-19 Collaboration, Anne Elmer, Nathalie Kingston, Barbara Graves, Emma Le Gresley, Daniela Caputo, Kenneth GC Smith, John R. Bradley, Lourdes Ceron-Gutierrez, Paulina Cortes-Acevedo, Gabriela Barcenas-Morales, Michelle Linterman, Laura McCoy, Chris Davis, Emma Thomson, Paul A. Lyons, Eoin McKinney, Rainer Doffinger, Mark Wills, Ravindra K. Gupta

**Affiliations:** Cambridge Institute of Therapeutic Immunology & Infectious Disease (CITIID), Cambridge, UK; Department of Medicine, University of Cambridge, Cambridge, UK; Division of Infection and Immunity, University College London, London, UK; The CITIID-NIHR BioResource COVID-19 Collaboration, see appendix 1 for author list; NIHR Cambridge Clinical Research Facility, Cambridge, UK; Department of Clinical Biochemistry and Immunology, Addenbrookes Hospital, UK; Laboratorio de Inmunologia, S-Cuautitlán, UNAM, Mexico; Babraham Institute, Cambridge, UK

**Keywords:** SARS-CoV-2, COVID-19, immune response, vaccine, neutralising antibodies, T cell, B cell repertoire

## Abstract

Two dose mRNA vaccination provides excellent protection against SARS-CoV-2. However, there are few data on vaccine efficacy in elderly individuals above the age of 80^1^. Additionally, new variants of concern (VOC) with reduced sensitivity to neutralising antibodies have raised fears for vulnerable groups. Here we assessed humoral and cellular immune responses following vaccination with mRNA vaccine BNT162b2^2^ in elderly participants prospectively recruited from the community and younger health care workers. Median age was 72 years and 51% were females amongst 140 participants. Neutralising antibody responses after the first vaccine dose diminished with increasing age, with a marked drop in participants over 80 years old. Sera from participants below and above 80 showed significantly lower neutralisation potency against B.1.1.7, B.1.351 and P.1. variants of concern as compared to wild type. Those over 80 were more likely to lack any neutralisation against VOC compared to younger participants following first dose. The adjusted odds ratio for inadequate neutralisation activity against the B.1.1.7, P.1 and B.1.351 variant in the older versus younger age group was 4.3 (95% CI 2.0-9.3, p<0.001), 6.7 (95% CI 1.7-26.3, p=0.008) and 1.7 (95% CI 0.5-5.7, p=0.41). Binding IgG and IgA antibodies were lower in the elderly, as was the frequency of SARS-CoV-2 Spike specific B-memory cells. We observed a trend towards lower somatic hypermutation in participants with suboptimal neutralisation, and elderly participants demonstrated clear reduction in class switched somatic hypermutation, driven by the IgA1/2 isotype. SARS-CoV-2 Spike specific T-cell IFNγ and IL-2 responses fell with increasing age, and both cytokines were secreted primarily by CD4 T cells. We conclude that the elderly are a high risk population that warrant specific measures in order to mitigate against vaccine failure, particularly where variants of concern are circulating.

## Background

Vaccines designed to elicit protective immune responses remain the key hope for containing the SARS-CoV-2 pandemic. In particular, mRNA vaccines have shown excellent efficacy using a two-dose approach, separated by a three or four week gap^2,3^. Although data on neutralising responses as a correlate of protection are increasing^4,5^, few data on neutralising responses or vaccine efficacy in elderly individuals above the age of 80 are available in trial settings^1^. This is even more pertinent for settings where a dosing interval of 12-16 weeks or even more has been implemented to maximise first dose administration^6^. In the absence of adequate clinical trial data, ‘real world’ data on vaccine responses are vital in order to understand the likely efficacy of vaccination using this dosing regime in those who are at greatest risk of severe disease and death^7^.

Additionally, the emergence of new variants with increased transmissibility^8^, reduced sensitivity to vaccine elicited antibodies^9^ and reduced clinical efficacy in preventing infection^10^ has raised fears for vulnerable groups where magnitude and quality of immune responses may be suboptimal. Here we assessed humoral and cellular immune responses following vaccination with mRNA-based vaccine BNT162b2^2^ in unselected elderly participants from the community and younger health care workers.

## Results

### Neutralisation of SARS-CoV-2 following mRNA vaccination in the elderly

One hundred and forty participants received at least one vaccination, with median age 72 years (IQR 44-83) and 51% of participants female (Extended Data Figure 1). We first validated the use of a pseudotyped virus (PV) system to investigate neutralisation, by comparing geometric mean titres (GMT) between PV expressing Wuhan-1 D614G spike and a B.1 lineage live virus isolate, using sera isolated from thirteen individuals after two vaccine doses (Extended data Figure 2a). We observed high correlation between the two approaches, consistent with previous literature^11^, and therefore proceeded with the PV system.

We explored the association between age and ability to neutralise virus by plotting the proportion of individuals who had detectable virus neutralisation after the first dose at a given age. This analysis showed a non-linear relationship with marked drop around the age of 80 (Figure 1a). Given this observation of a non-linear change in a correlate of protection we performed selected subsequent analyses with age both as a continuous variable and as a categorical variable. When individuals 80 years and above were tested between 3 and 12 weeks post first dose, around half had no evidence of neutralisation (Extended data Figure 2b). GMT was lower in participants 80 years and older than in younger individuals [48.2 (95% CI 34.6-67.1) vs 104.1 (95% CI 69.7-155.2) p<0.0001 Table 1, Figure 1b]. Geometric mean neutralisation titre (GMT) after the first dose (but not second dose) showed evidence of a modest inverse association with age (Extended data Figure 2c). The duration of the interval between first and second dose (three versus twelve weeks) did not appear to impact GMT following the second dose, though numbers were limited (Extended data Figure 2d). There was evidence for prior COVID-19 infection in 5 individuals in each group using a clinically accredited assay for N antibodies^9^ (Table 1), and this was adjusted for in multivariable analyses (Table 2, Supplementary Table 1). Sera from vaccinated individuals exhibited an increase in neutralising titres between the first and second doses for individuals both below and above the age of 80 years (Figure 1b). In those participants with suboptimal or no neutralisation who received second doses within the study period (examples shown in Figure 1c), testing after the second dose showed that all responded (Table 1 and Figure 1b).

**Table 1:**
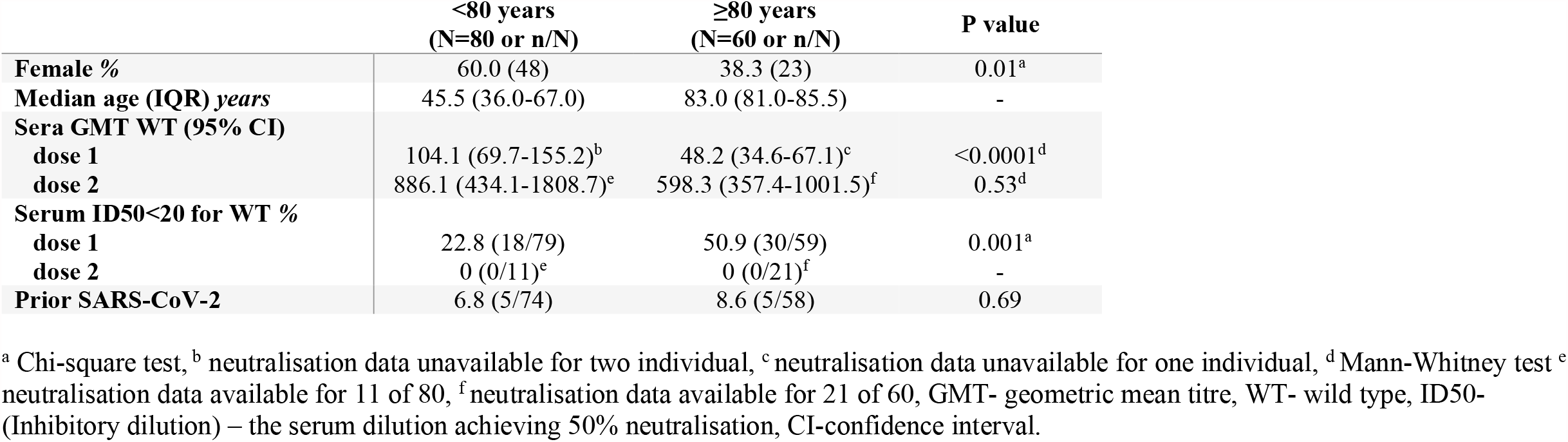
Characteristics of study participants.

|  | **<80 years**<br>**(N=80 or n/N)** | **≥80 years**<br>**(N=60 or n/N)** | **P value** |
| --- | --- | --- | --- |
| **Female %** | 60.0 (48) | 38.3 (23) | 0.01<sup>a</sup> |
| **Median age (IQR) years** | 45.5 (36.0-67.0) | 83.0 (81.0-85.5) | - |
| **Sera GMT WT (95% CI)** |  |  |  |
| **dose 1** | 104.1 (69.7-155.2)<sup>b</sup> | 48.2 (34.6-67.1)<sup>c</sup> | <0.0001<sup>d</sup> |
| **dose 2** | 886.1 (434.1-1808.7)<sup>e</sup> | 598.3 (357.4-1001.5)<sup>f</sup> | 0.53<sup>d</sup> |
| **Serum ID50<20 for WT %** |  |  |  |
| **dose 1** | 22.8 (18/79) | 50.9 (30/59) | 0.001<sup>a</sup> |
| **dose 2** | 0 (0/11)<sup>e</sup> | 0 (0/21)<sup>f</sup> | - |
| **Prior SARS-CoV-2** | 6.8 (5/74) | 8.6 (5/58) | 0.69 |

<sup>a</sup> Chi-square test, <sup>b</sup> neutralisation data unavailable for two individual, <sup>c</sup> neutralisation data unavailable for one individual, <sup>d</sup> Mann-Whitney test <sup>e</sup> neutralisation data available for 11 of 80, <sup>f</sup> neutralisation data available for 21 of 60, GMT- geometric mean titre, WT- wild type, ID50- (Inhibitory dilution) – the serum dilution achieving 50% neutralisation, CI-confidence interval.

**Table 2:** Neutralisation in participants after the first dose of BNT162b2 vaccine against wild type and B.1.1.7 spike mutant pseudotyped viruses.

|  | Number | Risk ID50<20 | Unadjusted OR<br>(95% CI) | P value | Adjusted OR* (95%<br>CI) | P value |
| --- | --- | --- | --- | --- | --- | --- |
| **WT** |  |  |  |  |  |  |
| **Age group years** |  |  |  |  |  |  |
| <80 | 78 | 23.1 (18/78) | 1 |  | 1 |  |
| ≥ 80 | 59 | 50.9 (30/59) | 3.4 (1.7-7.2) | 0.001 | 3.7 (1.7-8.1) | 0.001 |
| **Sex** |  |  |  |  |  |  |
| Male | 68 | 32.4 (22/68) | 1 |  | 1 |  |
| Female | 69 | 37.7 (26/69) | 1.2 (0.6- 2.6) | 0.52 | 1.4 (0.6-3.3) | 0.39 |
| **Time since dose 1 weeks** |  |  |  |  |  |  |
| 3-8 | 68 | 29.4 (20/68) | 1 |  | 1 |  |
| 9-12 | 69 | 40.6 (28/69) | 1.6 (0.8-3.3) | 0.17 | 1.6 (0.7-3.6) | 0.25 |
| **Previous COVID-19** |  |  |  |  |  |  |
| No | 121 | 35.5 (43/121) | 1 |  | 1 |  |
| Yes | 10 | 40.0 (4/100) | 1.2 (0.3-4.5) | 0.28 | 1.1 (0.3-4.6) | 0.92 |
| **B.1.1.7** |  |  |  |  |  |  |
| **Age group years** |  |  |  |  |  |  |
| <80 | 77 | 25.9 (20/77) | 1 |  | 1 |  |
| ≥ 80 | 58 | 60.3 (35/58) | 4.3 (2.1-9.0) | <0.001 | 4.3 (2.0-9.3) | <0.001 |
| **Sex** |  |  |  |  |  |  |
| Male | 67 | 43.3 (29/67) | 1 |  | 1 |  |
| Female | 68 | 38.2 (26/68) | 0.8 (0.4-1.6) | 0.55 | 1.2 (0.6-2.8) | 0.59 |
| **Time since dose 1 weeks** |  |  |  |  |  |  |
| 3-8 | 66 | 42.4 (28/66) | 1 |  | 1 |  |
| 9-12 | 69 | 39.1 (27/69) | 0.9 (0.4-1.7) | 0.70 | 0.7 (0.3-1.6) | 0.41 |
| **Previous COVID-19** |  |  |  |  |  |  |
| No | 120 | 41.7 (50/120) | 1 |  | 1 |  |
| Yes | 10 | 40.0 (4/10) | 0.9 (0.3-3.5) | 0.92 | 0.9 (0.2-3.6) | 0.88 |

\* Mutually adjusted for other variables in the table. WT- wild type, B.1.1.7- Spike mutant with N501Y, A570D, ΔH69/V70, Δ144/145, P681H, T716I, S982A and D1118H, OR- odds ratio, ID50- (Inhibitory dilution) – the serum dilution achieving 50% neutralisation, ns- non-significant, CI-confidence interval.

**Figure 1.**
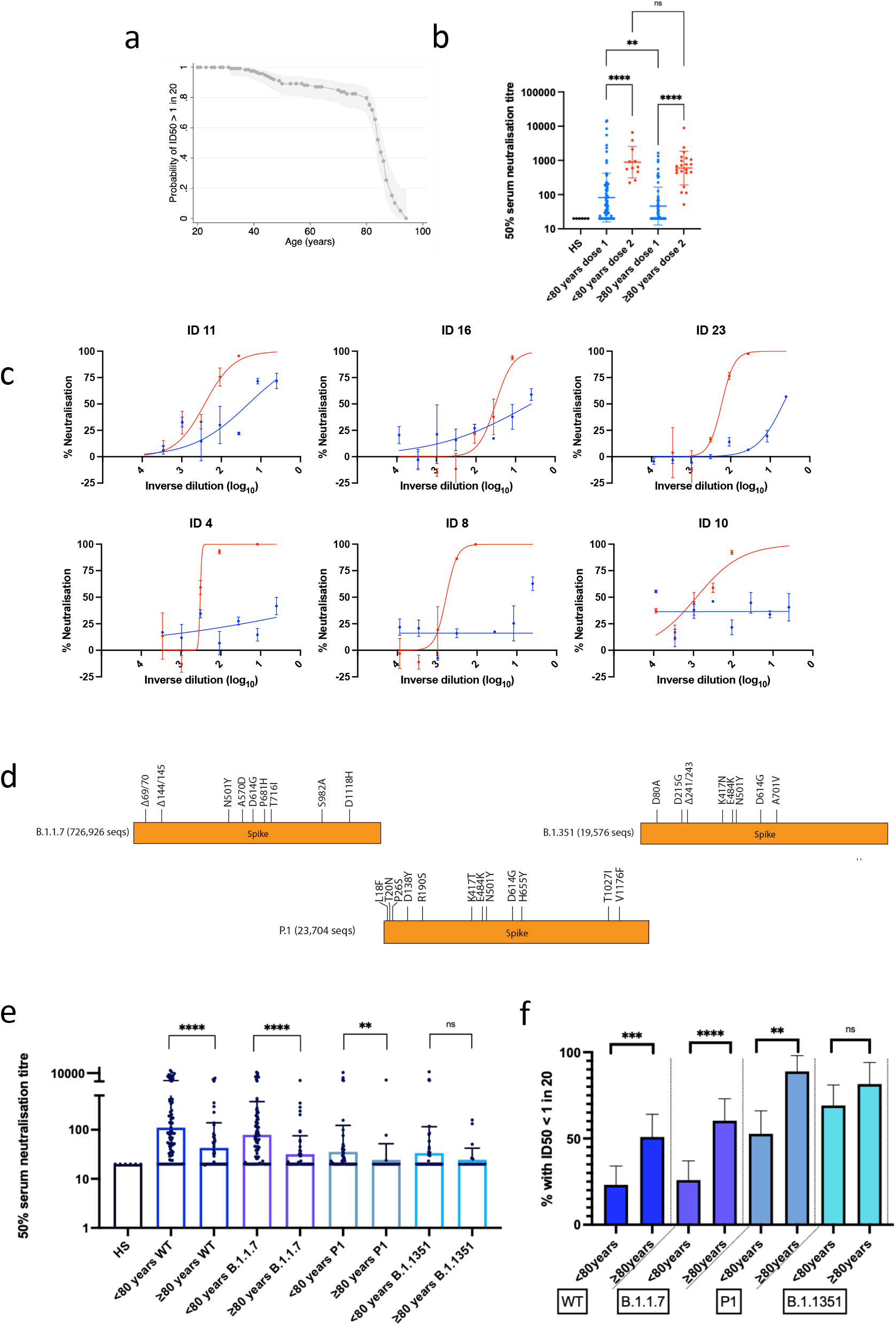
SARS-CoV-2 neutralisation by Pfizer BNT162b2 vaccine sera. **a**. Proportion of individuals with detectable serum neutralisation of PV after the first dose of Pfizer BNT162b2 vaccine by age. Cut off for serum neutralisation is the inhibitory dilution at which 50% inhibition of infection is achieved, ID50 of 20. The probability is bound by 95% confidence interval. **b**. SARS-CoV-2 PV neutralisation by Pfizer BNT162b2 first and second dose vaccine sera. Data are shown as mean ID50 values for individuals after Dose 1 (n=138) and after Dose 2 (n=32). Geometric mean with s.d is shown. Each point is a mean of technical replicates from two experiment repeats. **c**. Serum neutralisation of PV after Dose 1 (blue) or dose 2 (red) by age-group <80 years (n=79), ≥80 years (n=59). **c**. Neutralisation curves for serum from six individuals with reduced responses after first dose (blue) and increased neutralization activity after second dose (red) of Pfizer BNT162b2 vaccine against pseudovirus expressing wild type Spike (D614G). Neutralisation curves are means of technical replicates, plotted with error bars representing standard error of mean.**d**. diagram depicting spike mutations in variants of concern, along with number of sequences in GISAID **e**. Impact of SARS-CoV-2 variants of concern on neutralisation by Pfizer BNT162b2 dose 1 vaccine sera. WT (n=138), B.1.1.7 (n=135) Spike mutant B.1.351 (n=82) Spike mutant, P.1 (n=82). Geometric mean titre and s.d are shown. **f**. The proportion of participant vaccine sera with undetectable neutralistion of WT and Spike mutant (ID50 < 1 in 20 dilution of sera). WT (n=138), B.1.1.7 (n=135) Spike mutant; B.1.351 (n=82) spike mutant, P.1 (n=82) spike mutant. GMT with s.d are representative of two independent experiments each with two technical repeats. Mann-Whitney test was used for unpaired comparisons and Wilcoxon matched-pairs signed rank test for paired comparisons. p-values * <0.05, ** <0.01, **** <0.0001, ns not significant, HS – human AB serum control, r– Pearson’s correlation coefficient, β- slope/regression coefficient, p p-value. Bonferroni adjustment was made for multiple comparisons in linear regression.

### Neutralisation of SARS-CoV-2 variants of concern by vaccine elicited sera in the elderly

Given our observation that the participants 80 years old and older had lower neutralisation responses following first dose, we hypothesised that this could lead to sub protective neutralising responses against B.1.1.7, B.1.351 and P.1 variants of concern (VOC), originating in UK, South Africa and Brazil respectively (Figure 1d). We therefore examined serum neutralisation by age group against PV bearing WT or the three VOC spike proteins (Figure 1e, f). There was a clear reduction in neutralising titres against VOC as previously noted (Figure 1e), with the over 80 year olds exhibiting lower titres than younger individuals for all except B.1.351 where neutralisation was very poor universally following the first dose. When we analysed the proportions of individuals with no detectable neutralisation we observed the same pattern (Figure 1f). The adjusted odds ratio for achieving inadequate neutralisation against WT was 3.7 (95% CI 1.7-8.1, p 0.001) for participants 80 years and older versus those younger than 80 years (Table 2). The adjusted odds ratio for inadequate neutralisation activity against the B.1.1.7, P.1 and B.1.351 variant in the older versus younger age group was 4.3 (95% CI 2.0-9.3, p<0.001), 6.7 (95% CI 1.7-26.3, p=0.008) and 1.7 (95% CI 0.5-5.7, p=0.41) respectively (Table 2 and Supplementary Table 1).

### Binding Antibody responses and B cell Repertoire Analyses

Binding antibody responses to full length WT Wuhan-1 Spike were comprehensively measured using a clinically accredited particle based assay that we have previously described^9^. IgG and all IgG subclasses against Spike increased between vaccine doses (Figure 2a), reaching levels after the second dose similar to those observed following natural infection. IgG against Spike declined with age mirroring the neutralisation titres (Figure 2b, Extended Data Figure 3a). The concentration of total and subclass anti-Spike IgGs were significantly lower in the 80 years and older age group (Figure 2c). IgG and subclasses showed correlation with neutralisation (Figure 2c and Extended Data Figure 3b). IgA responses were detected both in convalescent sera (from individuals hospitalised in early-mid 2020) and after both doses, with an increase between the two time points (Extended Data Figure 3c). Spike specific IgA also correlated with neutralisation after dose 1(Extended Data Figure 3d). In addition, PBMC phenotyping by flow cytometry revealed neutralisation in the over 80 age group was associated with higher proportion of spike specific IgG+ IgM-CD19+ B memory cells (Figure 3e). Interestingly this did not differentiate neutralisers from non-neutralisers in the under 80 group (Figure 3e, Extended Data Figure 4b).

**Figure 2:**
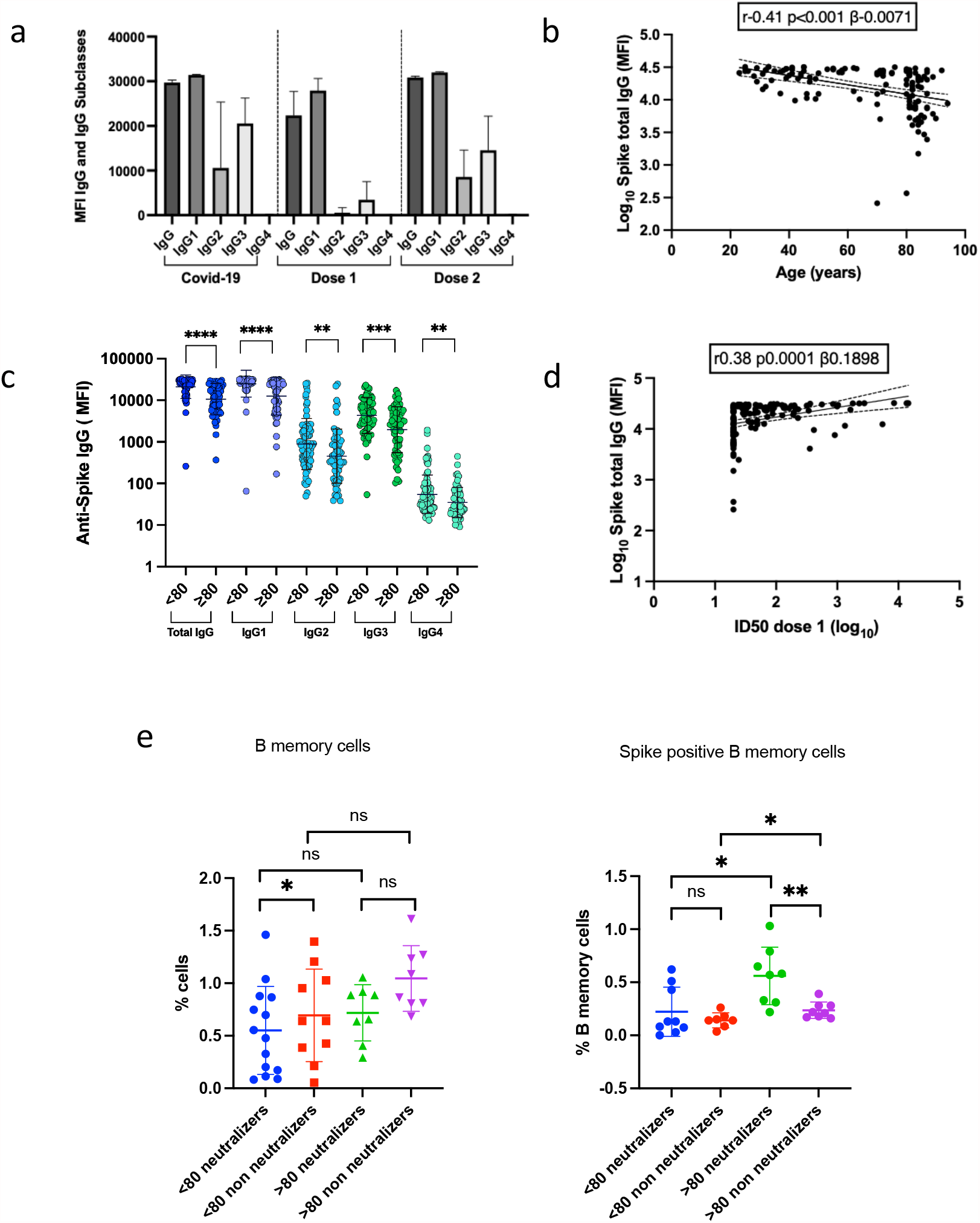
SARS-CoV-2 spike binding antibody responses and SARS-CoV-2 spike specific B memory cells in blood following vaccination with Pfizer BNT162b2 vaccine. **a**. Anti-Spike IgG-total and subclasses after first and second dose of vaccine compared to individuals with prior infection. **b**. Correlation between anti-Spike IgG binding antibody responses after first dose vaccine and age (n=134). **c**. Anti-Spike IgG subclass responses to first dose vaccine stratified by age <80 and ≥80 years. **d**. Correlations between anti-Spike IgG (n=134) binding antibody responses and neutralisation by vaccine sera against SARS-CoV-2 in a spike lentiviral pseudotyping assay expressing wild type Spike (D614G). **e**. CD19+ B memory (as % of PBMC) and SARS-CoV-2 spike specific B memory CD19+ IgG+ IgM-cells (as % of memory B cells) from FACS sorted PBMC. (n=16 above 80 and n=16 below 80 stratified by neutralizing response after first dose, n=8 in each category) MFI – mean fluorescence intensity. S – Spike, N – nucleocapsid, RBD – Spike receptor binding domain. Mann-whitney test was used for unpaired comparisons. p-values * <0.05, **<0.01, *** <0.001, ****<0.0001, ns-not significant HS – human AB serum control, Scatter plots show linear correlation line bounded by 95% confidence interval, r– Pearson’s correlation coefficient, p-P value, β slope/regression coefficient.

**Figure 3:**
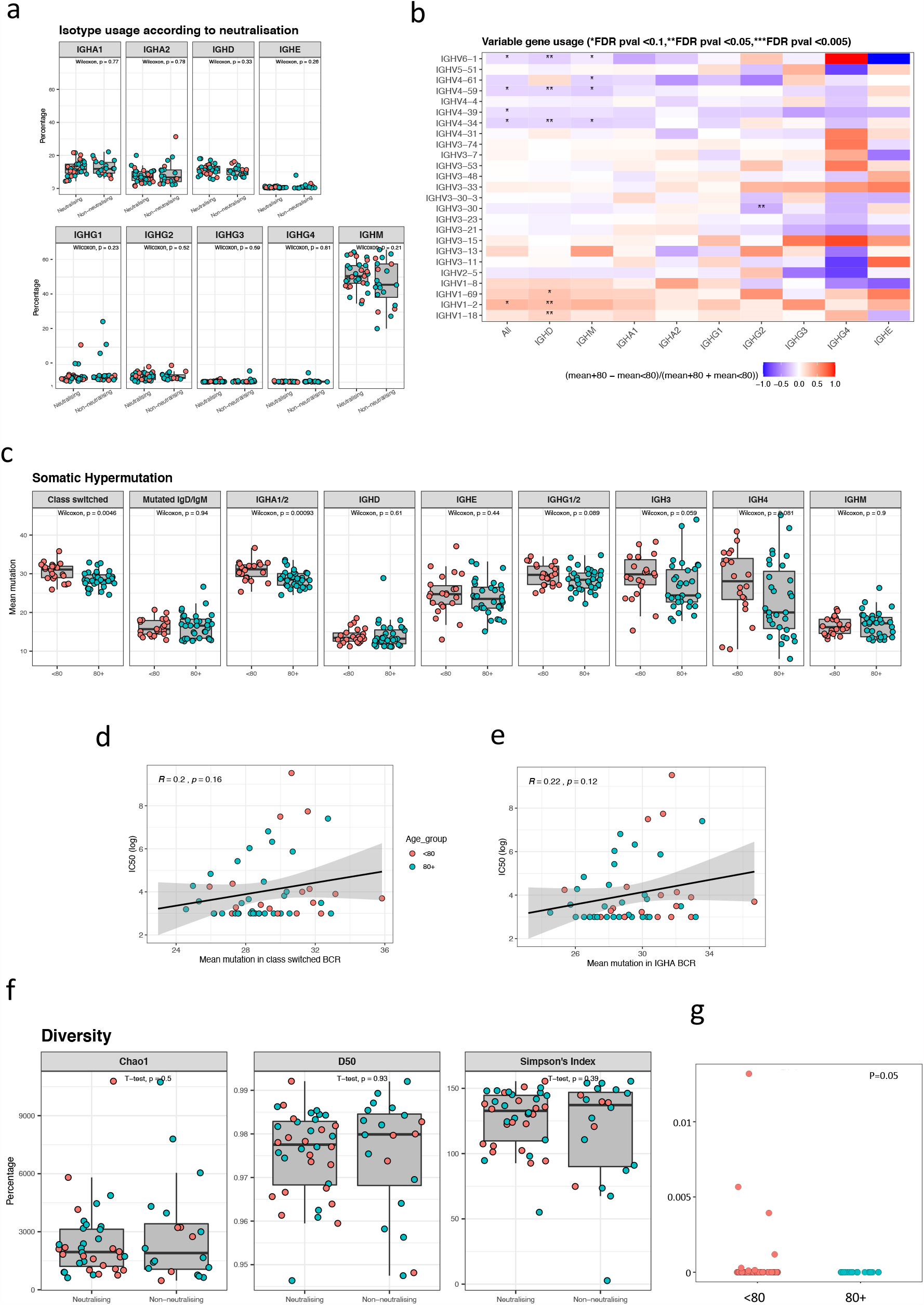
B cell repertoire following vaccination with first dose of Pfizer BNT162b2 vaccine. **a**. Boxplots showing Isotype usage according to unique VDJ sequence comparing participants <80 vs > 80 years old and association with neutralisation of spike pseudotyped virus. Neutralisation cut-off for 50% neutralisation was set at 20. **b**. Heat map showing V gene usage, comparing under 80 year olds with 80 year olds and older. A Benjamini Hochberg FDR correction was used, setting the threshold at 0.1. **B cell somatic hypermutation and BCR diversity following vaccination with first dose of Pfizer BNT162b2 vaccine. c**. Boxplots showing mean somatic hypermutation comparing <80 year olds with > 80 year old participants, grouped according to isotype class **d**. Correlation between somatic hypermutation in class-switched isotypes and IC50. **e**. Correlation between somatic hypermutation in IgHA BCR and IC50. **f**. Diversity Indices. The inverse is depicted for the Simpson’s index is normalised. **g**. BCR comparison with public clones known to be associated with SARS-CoV-2 neutralisation via the CoV-AbDab database (Raybould et al., 2020). Convergent clones were annotated with the same IGHV and IGHJ segments, had the same CDR-H3 region length and were clustered based on 85% CDR-H3 sequence amino acid homology. A cluster was considered convergent with the CoV-AbDab database if it contained sequences from post-vaccinated individuals and from the database.

Next B cell Repertoire (BCR) sequencing on bulk PBMCs was performed to assess isotype and variable gene usage, somatic hypermutation and diversity of the repertoire between the two age groups and in relation to neutralisation. All patients analysed had been vaccinated at a minimum 17 days prior. After correcting for multiple comparisons, there were no differences in isotype proportions between the two age groups (Extended Data Figure 5), or by neutralisation (Figure 3a). We next looked for skewing in V gene use (Figure 3b). We found an increase in usage of the IGHV4 family in the older age group with an increased proportion of IGHV4.34, IGHV4.39, IGHV4.59 and IGHV4.61 whilst in the younger age group there was an increase in usage of the IGHV1 family with increases in IGHV1.18 and IGHV1.69D. We did not find any significant differences in V gene usage with neutralisation (Extended Data Figure 5).

Differences in somatic hypermutation could impact neutralisation through antibody affinity maturation. We found that participants 80 years or older had a lower level of somatic hypermutation compared with the under 80 year olds in class-switched BCRs, which was driven by the IgA1/2 isotype (Figure 3c). There appeared to be a correlation between IC50 and degree of somatic hypermutation in class-switched BCRs (Figure 3d) driven predominantly by IGHA (Figure 3e). We did not observe a relationship between titres of IgG and mutation in BCRs for IgG1/2 (Extended Data Figure 5). To assess whether there was greater clonal expansion in those younger than 80 years old that might explain higher neutralising responses, we calculated richness, using the Chaol1 measure and diversity using D50, Simpson’s and Shannon-weiner indices. We did not find any significant differences between age groups or a relationship between measures of diversity and neutralisation potency (Figure 3f, Extended Data Figure 5).

We next examined the BCR for public clones known to be associated with SARS-CoV-2 neutralisation. We explored the convergence between BCR clones present in our study with the CoV-AbDab database, a resource detailing all published antibodies shown to bind SARS-CoV-2^12^. Convergent clones were annotated with the same IGHV and IGHJ segments, had the same CDR-H3 region length and were clustered based on 85% CDR-H3 sequence amino acid homology. A cluster was considered convergent with the CoV-AbDab database if it contained sequences from post-vaccinated individuals and from the database. This analysis revealed that participants under 80 had a higher frequency of convergent clones in keeping with increased neutralization when compared with the >80 age group (Figure 3g).

### T cell responses to SARS-CoV-2 spike following mRNA vaccination

Whilst neutralising antibodies are increasingly recognised as dominating protection against initial infection^4,13^, T cells may also play a role where neutralising antibody titres are low^14^, possibly limiting disease progression^5^. We therefore determined the T cell response to SARS-CoV-2 spike protein in vaccinees by stimulating PBMC with overlapping peptide pools to the wild type SARS-CoV-2 spike, using IFNγ and IL-2 FluoroSpot assay to enumerate spike specific T cells. With the same peptide pool, we also stimulated i. PBMC that had been collected and biobanked between 2014-2016 - representing a healthy SARS-CoV-2 unexposed population to provide a background control, ii. PBMC from donors who had RT-PCR confirmed infection with SARS-CoV-2 for a comparison of T cell responses following natural infection. When we plotted IFNγ spike specific T cell responses against age as a continuous variable there was a negative correlation and drop off at around 80 years (Figure 4a). A similar though less pronounced effect was seen for IL-2 (Figure 4b). However, there did not appear to be a relationship between cytokine production by PBMC and neutralisation titre following first dose (Extended Data Figure 6).

**Figure 4:**
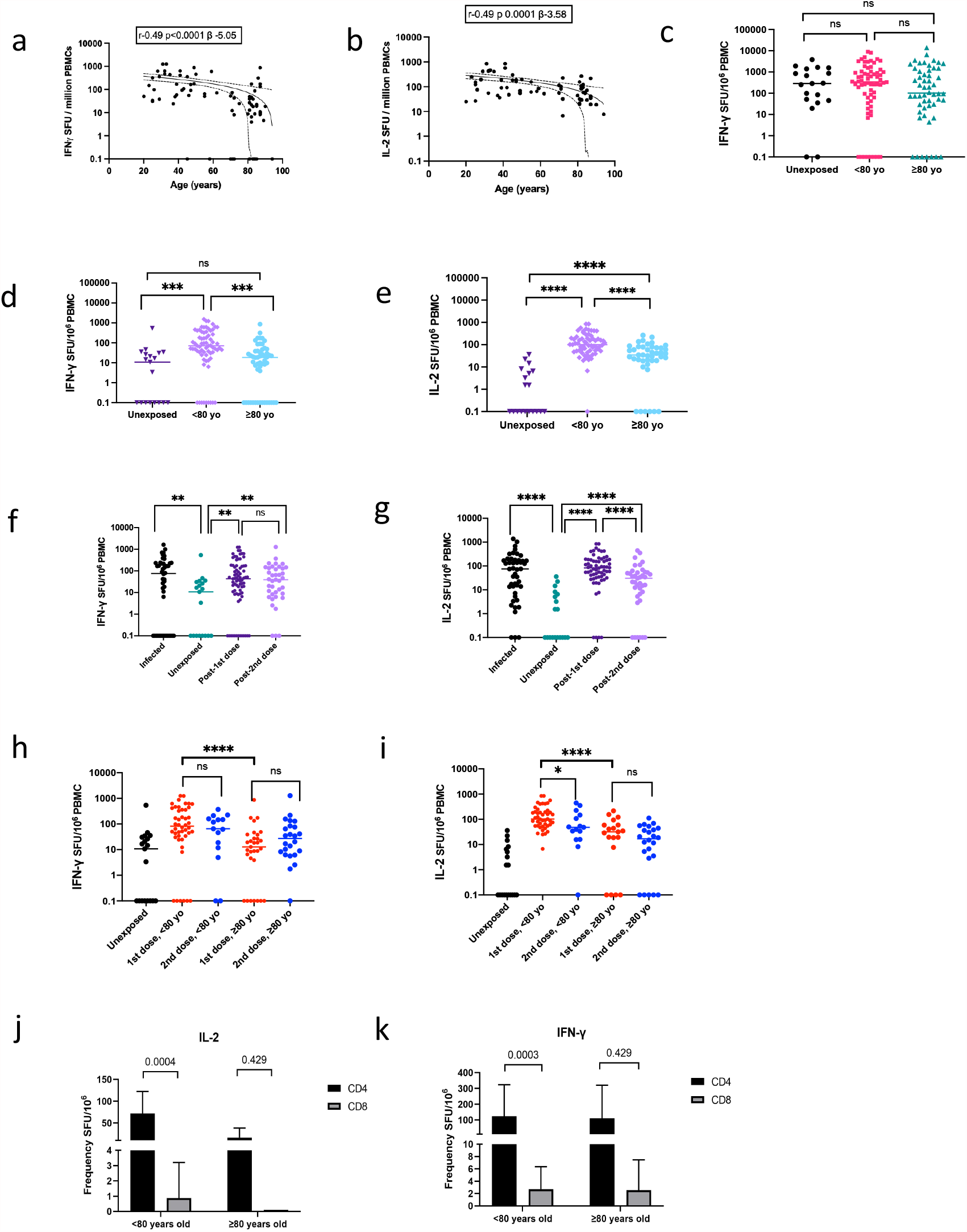
T cell responses to Pfizer BNT162b2 vaccine after the first and second doses of vaccine. FluoroSpot analysis by age for **a**. IFNγ and **b**. IL-2 T cell responses specific to SARS-CoV-2 Spike protein peptide pool following PBMC stimulation. **c**. FluoroSpot interferon gamma PBMC responses to peptide pool of Cytomegalovirus, Epstein Barr virus and Influenza virus (CEF) Response from unexposed stored PBMC 2014-2016 n=20), <80yo (n=46) and >80yo (n=35) three weeks after the first doses of Pfizer BNT162b2 vaccine. FluoroSpot analysis for **d**. IFNγ and **e**. IL-2 T cell responses specific to SARS-CoV-2 Spike protein peptide pool following PBMC stimulation of a cohort of unexposed (stored PBMC 2014-2016 n=20) and vaccinees <80yo IFNγ (n=46), IL-2 (n= 44) and >80yo IFNγ (n=35), IL-2 (n=27) three weeks or more after the first doses of Pfizer BNT162b2 vaccine. **f**. FluoroSpot analysis for IFNγ and **g**. IL-2 T cell responses specific to SARS-CoV-2 Spike protein peptide pool following PBMC stimulation of a cohort of infected (n=46), unexposed (n=20) and all vaccinees three weeks or more after the first doses IFNγ (n=77), IL-2 (n=64) and three weeks after second IFNγ and IL-2 (n=39) of Pfizer BNT162b2 vaccine. **h**. FluoroSpot analysis for IFNγ and **i**. IL-2 T cell responses specific to SARS-CoV-2 Spike protein peptide pool following PBMC stimulation of a cohort of unexposed (n=20) and vaccinees <80yo IFNγ (n=46), IL-2 (n=45) and >80yo IFNγ (n=31), IL-2 (n=19) three weeks after the first doses and <80yo IFNγ (n=15), IL-2 (n=15) and >80yo IFNγ (n=24), IL-2 (n=24) three weeks after second dose of Pfizer BNT162b2 vaccine. FluoroSpot analysis for **j**. IFNγ and **k**. IL-2 CD4 and CD8 T cell responses specific to SARS-CoV-2 Spike protein peptide pool following stimulation following column based PBMC separation. Mann-whitney test was used for unpaired comparisons and Wilcoxon matched-pairs signed rank test for paired comparisons. p-values * <0.05, ns not significant Mann-whitney test was used for unpaired comparisons and Wilcoxon matched-pairs signed rank test for paired comparisons. p-values ** <0.01, *** <0.001, ns not significant.

Following the first dose of vaccine, the frequency of IFNγ secreting T cells against a peptide pool including Cytomegalovirus, EBV and Flu (CEF+) specific peptides did not differ by age category and was similar to healthy SARS-CoV-2 unexposed controls (Figure 4c). This indicates that differences in observed responses were likely vaccine specific and unlikely due to generalised suboptimal T cell responses/immune paresis. However, IFNγ spike specific T cell responses were significantly larger than responses observed in an unexposed population for individuals under 80 (Figure 4d). Importantly however, in the 80 years and older participants, the IFNγ spike specific T cell responses were not different from the unexposed controls following first dose (Figure 4d). By contrast, spike specific IL-2 T cell frequencies were significantly greater than unexposed control in both age groups (Figure 4e). Interestingly, it appeared that whilst spike specific IFNγ and IL2 responses in PBMC were similar to those found after natural infection (Figure 4f,g), the second dose did not increase these responses, either overall (Figure 4f,g) or within age categories (Figure 4h,i).

We separated T cell subsets to further clarify the cells producing IFNγ and IL-2. PBMC from a sample of individuals <80 years and >80 years were depleted of CD4 or CD8 T cells and stimulated with spike peptide pool as before. Our results showed that the majority of IFNγ and IL-2 production was from the CD4+ T cells in vaccinated individuals (Figure 4j,k). The over 80 year olds had markedly lower spike specific IL2 CD4 T cell responses than their younger counterparts (Figure 4i).

CMV serostatus has been associated with poorer responses to vaccination and infections^15,16^ and we hypothesised that older individuals were more likely to be CMV positive and therefore poorer associated T cell responses to mRNA vaccination. We therefore ascertained CMV serostatus by ELISA testing and related this to T cell responses and serum neutralisation. As expected, CMV IgG positivity was higher in the 80 years and older age group (Extended Data Figure 7). Surprisingly however, CMV positive individuals in the 80 years and older group had significantly higher IFNγ, but not IL2, responses to SARS-CoV-2 spike peptides compared to the under 80 year old group (Extended Data Figure 7).

### Autoantibodies and inflammatory chemo/cytokines and responses to mRNA vaccination

Finally, we investigated possible interplay between senescence and mRNA vaccine responses. Autoantibodies and inflammatory cyto/chemokines are associated with immune senescence^17^. We first measured a panel of autoantibodies in the sera of 101 participants following the first dose of the BNT162b2 vaccine. 8 participants had positive autoantibodies for anti-myeloperoxidase (anti-MPO), 2 for anti-fibrillarin and 1 for anti-cardiolipin antibodies (Extended Data Figure 8). As expected, all but one of the participants with anti-myeloperoxidase autoantibody was over the age of 80 years (Extended Data Figure 8). There was a trend towards reduced anti-Spike IgG levels and serum neutralisation against wild type and B.1.17 Spike mutant in participants with positive autoantibodies, although this did not reach statistical significance, likely due to small numbers (Extended Data Figure 8). Next, we explored the association between serum cytokines/chemokines and neutralisation of SARS-CoV-2 PV as well as their association with age. PIDF, a known SASP (senescence associated secretory phenotype) molecule was the only molecule enriched in sera from participants over 80 years, and there was no association between any of these molecules and ability of sera to neutralise SARS-CoV-2 PV (Extended Data Figure 8).

## Discussion

Immune senescence is a well described phenomenon whereby responses to pathogens^18^ and indeed vaccines are impaired/dysregulated with age^19^. As an example, effective seasonal influenza vaccination of the elderly is a significant public health challenge due to greater morbidity and mortality in this group. Lower neutralizing antibody titres using standard dose influenza vaccines in elderly individuals has been addressed by using higher dose vaccine, highlighting that understanding of age related heterogeneity in vaccine responses can lead to policy change and mitigation ^20^.

Neutralising antibodies are likely the strongest correlate of protection from SARS-CoV-2 infection, as shown by vaccine efficacy studies, animal studies in mice and non-human primates, and data from early use of convalescent plasma in elderly patients ^4,5,13,14,21,22^. There is a lack of data on neutralising antibody immune responses following mRNA vaccination in the elderly and no data on variants of concern in this group. In a clinical study specifically looking at older adults vaccinated with BNT162b2 the GMT (geometric mean titre) after first dose was 12 in a set of 12 subjects between ages of 65 and 85 years, rising to 149 seven days after the second dose ^1^. Furthermore, in the Moderna 1273 mRNA vaccine study in older individuals (above 55 years), neutralisation was only detectable after the second dose, whilst binding antibodies were detectable after both doses^23^. In a randomised phase 1 study on BNT162b1 in younger (18-55) and older adults (65-85), Li et al observed lower virus neutralisation in the older age group at day 22 post first dose ^24^. These data parallel those in aged mice where ChAdOx nCov-19 vaccine responses were reported as being lower as compared to younger mice, and this was overcome by booster dosing^25^.

Here, in a substantial cohort of 140 individuals, we have shown not only an inverse relationship between age and neutralising responses following first dose of BNT162b2, but a more precipitous decline around the age of 80. Individuals 80 and above were prioritised for vaccination in the UK and elsewhere as they represented the group at greatest risk of severe COVID-19^26^. We found that around half of those above the age of 80 have a suboptimal neutralising antibody response after first dose vaccination with BNT162b2, accompanied by lower T cell responses compared to younger individuals. Individuals over 80 differed in four main respects that could explain poorer neutralisation of SARS-CoV-2. Firstly, serum IgG levels were lower, accompanied by a lower proportion of peripheral spike specific IgG+ IgM-CD19+ B memory cells. Secondly, the elderly displayed lower SHM in the BCR gene. Thirdly, the elderly had lower enrichment for public BCR clonotypes that are associated with neutralisation. Fourthly, the elderly displayed a marked reduction in IL-2 producing spike reactive CD4 T cells. Therefore possible explanations for poorer neutralising responses include lower concentrations (quantity) and/or lower affinity antibodies (quality) due to B cell selection, CD4 T cell help, or a combination of both.

Critically, we show that elderly individuals are likely at greater risk from VOC. When B.1.1.7 and P.1 variant spike PV were tested against sera in this study, a greater proportion of individuals in the over 80 age group lost all neutralising activity following first dose as compared to the wild type. As expected, B.1.351, known to have the highest degree of resistance to NAb^27^, did not appear to have preferential impact on the elderly after one dose, because sera from all ages had poor activity. The lack of neutralising activity against VOC following first dose in the elderly is of great concern in the current climate where variants are expanding globally and countries are struggling to acquire adequate vaccine supplies to ensure timely administration of both doses. We observed robust neutralising responses across all age groups after the second dose.

Following the second dose, binding IgG and IgA antibodies to Spike increased, mirroring levels seen in natural infection. Consistent with these data, the UK REACT study, a large, observational community based study, has shown that the prevalence of IgG positivity was 34.7% 21 days after the first dose of BNT162b2 in those over 80 years, increasing to 87.8% after the second dose^28^.

Although neutralising antibody responses appear critical, studies in rhesus macaques also show that CD8 T cells may play a role in contributing to protection from SARS-CoV-2 disease when neutralising antibody levels are low^14^. We speculate that this may explain why recent observational studies have demonstrated some effect against hospitalisation after 1 dose of the BNT162b2 or ChAdOx1nCoV-19 in the context of VOC ^10,29-32^. SARS-CoV-2 infection generates robust T cell responses to spike protein in the majority of individuals post infection. The IL2 response is predominantly produced by CD4+ T cells and that IFNγ production was seen in both spike specific CD4 and CD8+ T cell subsets^33,34^. By contrast, and in agreement with Anderson et al for the 1273 mRNA vaccine^23^, we found that spike specific IFNγ and IL-2 T cell responses to BNT162b2 mRNA vaccine were largely CD4 T cell derived. Phase I/II clinical trials of adenovirus vectored SARS-COV-2 vaccines have similarly showed a Th1 skewed response with elevated TNF*α*, IL2 and IFNγ being secreted by both CD4+ and CD8+ T cells^23,35-38^.

We found that both IFNγ and IL-2 PBMC responses were significantly lower in the over 80 age group and did not increase after the second dose, similar to findings for ChAdOx1^39^. Parry et al also reported suboptimal IFN γ responses in over 80 year olds following two doses of BNT162b2 (https://dx.doi.org/10.2139/ssrn.3816840). In particular it appeared that lower IL2 responses in PBMC may reflect very low spike specific CD4 IL2 responses in the over 80 age group.

SARS CoV-2 mRNA vaccination has been shown to generate potent T_FH_ and GC responses, correlating with neutralisation^40^. Interactions between B-cells and T follicular helper (T_FH_) in the germinal centre (GC) are needed for long-lived memory B cells/ plasma cells and high-affinity, class-switched antibodies^41,42^. In our B cell repertoire analyses we explored correlates for the impaired virus neutralisation by antibodies observed in a high proportion of older individuals. We found that participants >80 had a lower mean somatic hypermutation in class-switched BCRs, driven by the IgA1/2 isotype. There was a trend towards significant correlation between IC50 and somatic hypermutation in class-switched and in IGHA BCR.

Our data hint towards a mechanism whereby impaired T cell responses, as indicated by our data showing lower SARS-CoV-2 spike induced T cell cytokine production in the elderly, could impair generation of high affinity, class-switched, potently neutralising antibodies. In addition the lack of public BRC clones associated with neutralising antibodies in the elderly may relate to altered selection of B cell clones.

## Conclusion

*In vitro* virus neutralisation is increasingly recognised as the most significant correlate of protection from SARS-CoV-2 infection^4,5,13,14^. Whilst significant public health impact of vaccines is anticipated, and indeed has been demonstrated^43^, a significant proportion of individuals above 80 appear to require the second dose to achieve virus neutralisation and are disproportionately vulnerable to SARS-CoV-2 variants after the first dose. Our data caution against extending the dosing interval of BNT162b2 in the elderly population, particularly during periods of high transmission and risk from variants that are less susceptible to vaccine-elicited neutralising antibodies^44-47^. The mechanism of reduced neutralising responses in the elderly needs to be further delineated in order to inform mitigation strategies.

## Data Availability

Data are available on request from the corresponding author

## Acknowledgements

We would like to thank Cambridge University Hospitals NHS Trust Occupational Health Department. We would also like to thank the NIHR Cambridge Clinical Research Facility and staff at CUH, the Cambridge NIHR BRC Stratified Medicine Core Laboratory NGS Hub, the NIHR Cambridge BRC Phenotyping Hub, Petra Mlcochova, Steven A. Kemp, Martin Potts, Ben Krishna, Marianne Perera and Georgina Okecha. We would like to thank James Nathan, Leo James and John Briggs. RKG is supported by a Wellcome Trust Senior Fellowship in Clinical Science (WT108082AIA). DAC is supported by a Wellcome Trust Clinical PhD Research Fellowship. KGCS is the recipient of a Wellcome Investigator Award (200871/Z/16/Z). This research was supported by the National Institute for Health Research (NIHR) Cambridge Biomedical Research Centre, the Cambridge Clinical Trials Unit (CCTU), the NIHR BioResource and Addenbrooke’s Charitable Trust, the Evelyn Trust (20/75), UKRI COVID Immunology Consortium. GBM and PCA were supported by UNAM-FESC-PIAPI Program Code PIAPI2009 and by CONACyT 829997 fellowship. The views expressed are those of the authors and not necessarily those of the NIHR or the Department of Health and Social Care. IATMF is funded by a Sub-Saharan African Network for TB/HIV Research Excellence (SANTHE, a DELTAS Africa Initiative (grant DEL-15–006)) fellowship. We would like to thank Davide Corti for the VOC plasmids.

## Methods

### Study Design

Community participants or health care workers receiving the first dose of the BNT162b2 vaccine between the 14^th^ of December 2020 to the 10^th^ of February 2021 were consecutively recruited at Addenbrookes Hospital into the COVID-19 cohort of the NIHR Bioresource. Participants were followed up for up to 3 weeks after receiving their second dose of the BNT162b2 vaccine. They provided blood samples 3 to 12 weeks after their first dose and again 3 weeks after the second dose of the vaccine. Consecutive participants were eligible without exclusion. The exposure of interest was age, categorised into 2 exposure levels- < 80 and ≥ 80 years. The outcome of interest was inadequate vaccine-elicited serum antibody neutralisation activity at least 3 weeks after the first dose. This was measured as the dilution of serum required to inhibit infection by 50% (ID50) in an *in vitro* neutralisation assay. An ID50 of 20 or below was deemed as inadequate neutralisation. Binding antibody responses to Spike, Receptor binding domain and Nucleocapsid were measured by multiplex particle-based flow cytometry and Spike-specific T cell responses were measured by IFNy and IL-2 FLUOROSPOT assays. Measurement of serum autoantibodies and characterisation of the B cell receptor repertoire (BCR) following the first vaccine dose were exploratory outcomes.

We assumed a risk ratio of non-neutralisation in the ≥80 years group compared with <80 years group of 5. Using an alpha of 0.05 and power of 90% required a sample size of 50 with a 1:1 ratio in each group.

### Ethical approval

The study was approved by the East of England – Cambridge Central Research Ethics Committee (17/EE/0025). PBMC from unexposed volunteers previously recruited by the NIHR BioResource Centre Cambridge through the ARIA study (2014-2016), with ethical approval from the Cambridge Human Biology Research Ethics Committee (HBREC.2014.07) and currently North of Scotland Research Ethics Committee 1 (NS/17/0110).

### Statistical Analyses

Descriptive analyses of demographic and clinical data are presented as median and interquartile range (IQR) when continuous and as frequency and proportion (%) when categorical. The difference in continuous and categorical data were tested using Wilcoxon rank sum and Chi-square test respectively. Logistic regression was used to model the association between age group and neutralisation by vaccine-elicited antibodies after the first dose of the BNT162b2 vaccine. The effect of sex and time interval from vaccination to sampling as confounders were adjusted for. Linear regression was also used to explore the association between age as a continuous variable and log transformed ID50, binding antibody levels, antibody subclass levels and T cell response after dose 1 and dose 2 of the BNT162b2 vaccine. The time interval from vaccination to sampling was adjusted for. Bonferroni adjustment was made for multiple comparisons in the linear correlation analyses between binding antibody levels, ID50, age and T cell responses. The Pearson’s correlation coefficient for linear data and Spearman’s correlation for non-linear data was reported. Statistical analyses were done using Stata v13, Prism v9 and R (version 3.5.1).

### Generation of Mutants and pseudotyped viruses

Wild-type (WT) bearing 614G and B.1.1.7 bearing mutations del-69/70, del-144, N501Y, A570D, D614G, P681H, S982A, T716I and D1118H or K417N, E484K and N501Y pseudotyped viruses were generated as previously described^44^. In brief, amino acid substitutions were introduced into the D614G pCDNA_SARS-CoV-2_S plasmid as previously described^48^ using the QuikChange Lightening Site-Directed Mutagenesis kit, following the manufacturer’s instructions (Agilent Technologies, Inc., Santa Clara, CA). Sequences were verified by Sanger sequencing. The pseudoviruses were generated in a triple plasmid transfection system whereby the Spike expressing plasmid along with a lentviral packaging vector-p8.9 and luciferase expression vector-psCSFLW where transfected into 293T cells with Fugene HD transfection reagent (Promega). The viruses were harvested after 48 hours and stored at −80°C. TCID50 was determined by titration of the viruses on 293Ts expressing ACE-2 and TMPRSS2.

### Pseudotyped virus neutralisation assays

Spike pseudotype assays have been shown to have similar characteristics as neutralisation testing using fully infectious wild type SARS-CoV-2^11^. Virus neutralisation assays were performed on 293T cell transiently transfected with ACE2 and TMPRSS2 using SARS-CoV-2 Spike pseudotyped virus expressing luciferase^49^. Pseudotyped virus was incubated with serial dilution of heat inactivated human serum samples or sera from vaccinees in duplicate for 1h at 37°C. Virus and cell only controls were also included. Then, freshly trypsinized 293T ACE2/TMPRSS2 expressing cells were added to each well. Following 48h incubation in a 5% CO2 environment at 37°C, luminescence was measured using the Steady-Glo Luciferase assay system (Promega). Neutralization was calculated relative to virus only controls. Dilution curves were presented as a mean neutralization with standard error of the mean (SEM). 50% neutralization-ID50 values were calculated in GraphPad Prism. The limit of detection for 50% neutralisation was set at an ID50 of 20. The ID50 within groups were summarised as a geometric mean titre (GMT) and statistical comparison between groups were made with Mann-Whitney or Wilxocon ranked sign test.

### Live virus serum neutralisation assays

A549-Ace2-TMPRSS2 cells were seeded at a cell density of 2.4×10^4/well in a 96 well plate 24hrs before inoculation. Serum was titrated starting at a final 1:50 dilution with live B.1 virus PHE2 (EPI_ISL_407073) isolate being added at an MOI 0.01. The mixture was then incubating for 1hr prior to adding to the cells. 72hrs post infection the plates were fixed with 8% formaldehyde then stained with Coomassie blue for 30 minutes. The plates were washed and dried overnight before using a Celigo Imaging Cytometer (Nexcelom) to measure the staining intensity. Percentage cell survival was determined by comparing the intensity of the staining to an uninfected well. A non-linear sigmoidal 4PL model (Graphpad Prism 9) was used to determine the IC50 for each serum. The correlation between log transformed ID50 obtained from the pseudotyped virus and live virus systems were explored using linear regression. Pearson’s correlation coefficient was determined.

### SARS-CoV-2 serology by multiplex particle-based flow cytometry (Luminex)

Recombinant SARS-CoV-2 N, S and RBD were covalently coupled to distinct carboxylated bead sets (Luminex; Netherlands) to form a 3-plex and analyzed as previously described^50^. Specific binding was reported as mean fluorescence intensities (MFI).

### CMV serology

HCMV IgG levels determined using an IgG enzyme-linked immunosorbent (EIA) assay, HCMV Captia (Trinity Biotech, Didcot, UK) following manufacturer’s instructions, on plasma derived from clotted blood samples.

### Serum autoantibodies

Serum was screened for the presence of autoantibodies using the ProtoPlexTM autoimmune panel (Life Technologies) according to the manufacturer’s instructions. Briefly, 2.5μl of serum was incubated with Luminex MagPlex magnetic microspheres in a multiplex format conjugated to 19 full length human autoantigens (Cardiolipin, CENP B, H2a(F2A2) & H4 (F2A1), Jo-1, La/SS-B, Mi-2b, myeloperoxidase, proteinase-3, pyruvate dehydrogenase, RNP complex, Ro52/SS-A, Scl-34, Scl-70, Smith antigen, Thyroglobulin, Thyroid peroxidase, transglutaminase, U1-snRNP 68, whole histone) along with bovine serum albumin (BSA). Detection was undertaken using goat-anti-human IgG-RPE in a 96 well flat-bottomed plate and the plate was read in a Luminex xMAP 200 system. Raw fluorescence intensities (FI) were further processed in R (version 3.5.1) Non-specific BSA-bound FI was subtracted from background-corrected total FI for each antigen before log2 transformation and thresholding. Outlier values (Q3+1.5*IQR) in each distribution were defined as positive.

### Serum chemo/cytokine analysis

Serum proteins were quantified using a validated electro chemiluminescent sandwich assay (Mesoscale Discovery VPlex) quantification kit following the manufacturer’s instructions. Briefly both sera and standard calibration controls were incubated with SULFO-tagged antibodies targeting IFNγ, IL10, IL12p70, IL13, IL1β, IL2, IL4, IL6, IL8, TNFα, GC-CSF, IL1α, IL12, IL15, IL16, IL17A, IL5, IL7, TNFβ, VEGF, MCP1, MCP4, Eotaxin, Eotaxin3, IP10, MDC, MIP-1α, MIP-1β, TARC, IL17B, IL17C, IL17D, IL1RA, IL3, IL9, TSLP, VEGFA, VEGFC, VEGFD, VEGFR1/Flt1, PIGF, TIE2, FGF, ICAM1, VCAM1, SAA and CRP and read using an MSD MESO S600 instrument. Concentrations were calculated by comparison with an internal standard calibration curve fitted to a 4-parameter logistic model. Values below (19%) or above (0.0%) the reference range were imputed at the lower/upper limit of detection respectively. Association of each cytokine level with SARS-CoV-2 neutralising antibody titre, neutralisation status (1/0) and age was undertaken using Kendall’s Tau and Wilcoxon tests with FDR<5% considered significant.

### B Cell Receptor Repertoire Library Preparation

PBMC were lysed and RNA extracted using Qiagen AllPrep® DNA/RNA mini kits and Allprep® DNA/RNA Micro kits according to the manufactures protocol. The RNA was quantified using a Qubit. B cell receptor repertoire libraries were generated for 52 COVID-19 patients (58 samples) using as follows: 200ng of total RNA from PAXgenes (14ul volume) was combined with 1uL 10mM dNTP and 10uM reverse primer mix (2uL) and incubated for 5 min at 70°C. The mixture was immediately placed on ice for 1 minute and then subsequently combined with 1uL DTT (0.1 M), 1uL SuperScriptIV (Thermo Fisher Scientific), 4ul SSIV Buffer (Thermo Fisher Scientific) and 1uL RNAse inhibitor. The solution was incubated at 50 °C for 60 min followed by 15 min inactivation at 70 °C. cDNA was cleaned with AMPure XP beads and PCR-amplified with a 5’ V-gene multiplex primer mix and 3’ universal reverse primer using the KAPA protocol and the following thermal cycling conditions: 1cycle (95°C, 5min); 5cycles (98°C, 20s; 72°C, 30s); 5cycles (98°C, 15s; 65°C, 30s; 72°C, 30s); 19cycles (98 °C, 15s; 60°C, 30s; 72°C, 30s); 1 step (72°C, 5 min). Sequencing libraries were prepared using Illumina protocols and sequenced using 300-bp paired-end sequencing on a MiSeq machine.

### Sequence analysis

Raw reads were filtered for base quality using a median Phred score of ≥32 (http://sourceforge.net/projects/quasr/). Forward and reverse reads were merged where a minimum 20bp identical overlapping region was present. Sequences were retained where over 80% base sequence similarity was present between all sequences with the same barcode. The constant-region allele with highest sequence similarity was identified by 10-mer matching to the reference constant-region genes from the IMGT database. Sequences without complete reading frames and non-immunoglobulin sequences were removed and only reads with significant similarity to reference IGHV and J genes from the IMGT database using BLAST were retained. Immunoglobulin gene use and sequence annotation were performed in IMGT V-QUEST, and repertoire differences were performed by custom scripts in Python.

### Flow cytometry

The following antibodies or staining reagents were purchased from BioLegend: CD19 (SJ25C, 363028), CD3 (OKT3, 317328), CD11c (3.9, 301608), CD25 (M-A251, 356126), CD14 (M5E2,301836), and IgM (IgG1-k, 314524). CCR7 (150503, 561143) and IgG (G18-145, 561297) were obtained from BD Bioscience, CD45RA (T6D11, 130-113-359) from Miltyeni Biotech, and CD8a (SK1, 48-0087-42) from eBiosciences. The LIVE/DEAD™ Fixable Aqua Dead Cell Stain Kit was obtained from Invitrogen. Biotinylated Spike protein expressed and purified as previously described^51^ was conjugated to Streptavidin R-Phycoerythrin (PJRS25-1), or Streptavidin APC obtained from Agilent Technologies. PBMCs were isolated from study participants and stored in liquid nitrogen. Aliquots containing 10^7 cells were thawed and stained in PBS containing 2mM EDTA at 4 °C with the above antibody panel and then transferred to 0.04% BSA in PBS. Events were acquired on a FACSAria Fusion (BD Biosciences). Analyses were carried out in FlowJo version 10.7.1.

### IFNγ and IL2 FLUOROSpot T cell assays

Peripheral blood mononuclear cells (PBMC) were isolated from the heparinized blood samples using Histopaque-1077 (Sigma-Aldrich) and SepMate-50 tubes (Stemcell Technologies). Frozen PBMCs were rapidly thawed and diluted into 10ml of TexMACS media (Miltenyi Biotech), centrifuged and resuspended in 10ml of fresh media with 10U/ml DNase (Benzonase, Merck-Millipore via Sigma-Aldrich), PBMCs were then incubated at 37°C for 1h, followed by centrifugation and resuspension in fresh media supplemented with 5% Human AB serum (Sigma Aldrich) before being counted. PBMCs were stained with 2ul of LIVE/DEAD Fixable Far Red Dead Cell Stain Kit (Thermo Fisher Scientific) and live PBMC enumerated on the BD Accuri C6 flow cytometer.

### Overlapping Spike SARS-CoV-2 peptide stimulation

A peptide pool was generated using the following: 1. PepTivator SARS-CoV-2 Prot_S containing the sequence domains aa 304-338, 421-475, 492-519, 683-707, 741-770, 785-802, and 885 – 1273 and S1 N-terminal S1 domain of the surface glycoprotein (“S”) of SARS-Coronavirus 2 (GenBank MN908947.3, Protein QHD43416.1). 2. The PepTivator SARS-CoV-2 Prot_S1 containing the aa sequence 1–692. The peptides used are 15aa amino acids with 11 amino acid overlaps.

1.0 to 2.5 ⨯ 10^5^ PBMCs were incubated in pre-coated FluoroSpot^FLEX^ plates (anti IFNγand IL2 capture antibodies Mabtech AB, Nacka Strand, Sweden)) in duplicate with spike peptide pool mix as described above (specific for Wuhan-1, QHD43416.1) Spike SARS-CoV-2 protein (Miltenyi Biotech) or a mixture of peptides specific for Cytomegalovirus, Epstein Barr virus and Influenza virus (CEF+, (Miltenyi Biotech)) (final peptide concentration manufactures recommendation 1µg/ml/peptide, Miltenyi Biotech) in addition to an unstimulated (media only) and positive control mix (containing anti-CD3 (Mabtech AB) and Staphylococcus Enterotoxin B (SEB), (Sigma Aldrich)) at 37°C in a humidified CO2 atmosphere for 42 hours. The cells and medium were then decanted from the plate and the assay developed following the manufacturer’s instructions. Developed plates were read using an AID iSpot reader (Oxford Biosystems, Oxford, UK) and counted using AID EliSpot v7 software (Autoimmun Diagnostika GmbH, Strasberg, Germany). Peptide specific frequencies were calculated by subtracting for background cytokine specific spots (unstimulated control) and expressed as SFU/Million PBMC.

### CD4 and CD8 depletion from PBMC for subsequent FLUOROSpot analysis

Peripheral blood mononuclear cells were depleted of either CD4+ or CD8+ T cells by MACS using anti-CD4+ or anti-CD8+ direct beads (Miltenyi Biotec), according to manufacturer’s instructions, and separated by using an AutoMACS Pro (Miltenyi Biotec). Efficiency of depletion was determined by staining cells with a CD3-FITC, CD4-PE, and CD8-PerCPCy5.5 antibody mix (all BioLegend) and analyzed by flow cytometry.

## The CITIID-NIHR BioResource COVID-19 Collaboration

### Principal Investigators

Stephen Baker^2, 3^, Gordon Dougan^2, 3^, Christoph Hess^2,3,28,29^, Nathalie Kingston^22, 12^, Paul J. Lehner^2, 3^, Paul A. Lyons^2, 3^, Nicholas J. Matheson^2, 3^, Willem H. Owehand^22^, Caroline Saunders^21^, Charlotte Summers^,3,26,27,30^, James E.D. Thaventhiran^2, 3, 24^, Mark Toshner^3, 26, 27^, Michael P. Weekes^2^, Patrick Maxwell^22,30^, Ashley Shaw^30^

### CRF and Volunteer Research Nurses

Ashlea Bucke^21^, Jo Calder^21^, Laura Canna^21^, Jason Domingo^21^, Anne Elmer^21^, Stewart Fuller^21^, Julie Harris^43^, Sarah Hewitt^21^, Jane Kennet^21^, Sherly Jose^21^, Jenny Kourampa^21^, Anne Meadows^21^, Criona O’Brien^43^, Jane Price^21^, Cherry Publico^21^, Rebecca Rastall^21^, Carla Ribeiro^21^, Jane Rowlands^21^, Valentina Ruffolo^21^, Hugo Tordesillas^21^,

### Sample Logistics

Ben Bullman^2^, Benjamin J. Dunmore^3^, Stuart Fawke^32^, Stefan Gräf ^3,22,12^, Josh Hodgson^3^, Christopher Huang^3^, Kelvin Hunter^2, 3^, Emma Jones^31^, Ekaterina Legchenko^3^, Cecilia Matara^3^, Jennifer Martin^3^, Federica Mescia^2, 3^, Ciara O’Donnell^3^, Linda Pointon^3^, Nicole Pond^2, 3^, Joy Shih^3^, Rachel Sutcliffe^3^, Tobias Tilly^3^, Carmen Treacy^3^, Zhen Tong^3^, Jennifer Wood^3^, Marta Wylot^38^,

### Sample Processing and Data Acquisition

Laura Bergamaschi^2, 3^, Ariana Betancourt^2, 3^, Georgie Bower^2, 3^, Chiara Cossetti^2, 3^, Aloka De Sa^3^, Madeline Epping^2, 3^, Stuart Fawke^32^, Nick Gleadall^22^, Richard Grenfell^33^, Andrew Hinch^2,3^, Oisin Huhn^34^, Sarah Jackson^3^, Isobel Jarvis^3^, Ben Krishna3, Daniel Lewis^3^, Joe Marsden^3^, Francesca Nice^41^, Georgina Okecha^3^, Ommar Omarjee^3^, Marianne Perera^3^, Martin Potts^3^, Nathan Richoz^3^, Veronika Romashova^2,3^, Natalia Savinykh Yarkoni^3^, Rahul Sharma^3^, Luca Stefanucci^22^, Jonathan Stephens^22^, Mateusz Strezlecki^33^, Lori Turner^2, 3^,

### Clinical Data Collection

Eckart M.D.D. De Bie^3^, Katherine Bunclark^3^, Masa Josipovic^42^, Michael Mackay^3^, Federica Mescia^2,3^, Alice Michael^27^, Sabrina Rossi^37^, Mayurun Selvan^3^, Sarah Spencer^15^, Cissy Yong^37^

### Royal Papworth Hospital ICU

Ali Ansaripour^27^, Alice Michael^27^, Lucy Mwaura^27^, Caroline Patterson^27^, Gary Polwarth^27^

### Addenbrooke’s Hospital ICU

Petra Polgarova^30^, Giovanni di Stefano^30^

### Cambridge and Peterborough Foundation Trust

Codie Fahey^36^, Rachel Michel^36^

### ANPC and Centre for Molecular Medicine and Innovative Therapeutics

Sze-How Bong^23^, Jerome D. Coudert^35^, Elaine Holmes^39^

### NIHR BioResource

John Allison^22,12^, Helen Butcher^12,40^, Daniela Caputo^12,40^, Debbie Clapham-Riley^12,40^, Eleanor Dewhurst^12,40^, Anita Furlong^12,40^, Barbara Graves^12,40^, Jennifer Gray^12,40^, Tasmin Ivers^12,40^, Mary Kasanicki^12,30^, Emma Le Gresley^12,40^, Rachel Linger^12,40^, Sarah Meloy^12,40^, Francesca Muldoon^12,40^, Nigel Ovington^22,12^, Sofia Papadia^12,40^, Isabel Phelan^12,40^, Hannah Stark^12,40^, Kathleen E Stirrups^22,12^, Paul Townsend^22,12^, Neil Walker^22,12^, Jennifer Webster^12,40^.

21. Cambridge Clinical Research Centre, NIHR Clinical Research Facility, Cambridge University Hospitals NHS Foundation Trust, Addenbrooke’s Hospital, Cambridge CB2 0QQ, UK

22. University of Cambridge, Cambridge Biomedical Campus, Cambridge CB2 0QQ, UK

23. Australian National Phenome Centre, Murdoch University, Murdoch, Western Australia WA 6150, Australia

24. MRC Toxicology Unit, School of Biological Sciences, University of Cambridge, Cambridge CB2 1QR, UK

25. R&D Department, Hycult Biotech, 5405 PD Uden, The Netherlands

26. Heart and Lung Research Institute, Cambridge Biomedical Campus, Cambridge CB2 0QQ, UK

27. Royal Papworth Hospital NHS Foundation Trust, Cambridge Biomedical Campus, Cambridge CB2 0QQ, UK

28. Department of Biomedicine, University and University Hospital Basel, 4031Basel, Switzerland

29. Botnar Research Centre for Child Health (BRCCH) University Basel & ETH Zurich, 4058 Basel, Switzerland

30. Addenbrooke’s Hospital, Cambridge CB2 0QQ, UK

31. Department of Veterinary Medicine, Madingley Road, Cambridge, CB3 0ES, UK

32. Cambridge Institute for Medical Research, Cambridge Biomedical Campus, Cambridge CB2 0XY, UK

33. Cancer Research UK, Cambridge Institute, University of Cambridge CB2 0RE, UK

34. Department of Obstetrics & Gynaecology, The Rosie Maternity Hospital, Robinson Way, Cambridge CB2 0SW, UK

35. Centre for Molecular Medicine and Innovative Therapeutics, Health Futures Institute, Murdoch University, Perth, WA, Australia

36. Cambridge and Peterborough Foundation Trust, Fulbourn Hospital, Fulbourn, Cambridge CB21 5EF, UK

37. Department of Surgery, Addenbrooke’s Hospital, Cambridge CB2 0QQ, UK

38. Department of Biochemistry, University of Cambridge, Cambridge, CB2 1QW, UK

39. Centre of Computational and Systems Medicine, Health Futures Institute, Murdoch University, Harry Perkins Building, Perth, WA 6150, Australia

40. Department of Public Health and Primary Care, School of Clinical Medicine, University of Cambridge, Cambridge Biomedical Campus, Cambridge, UK

## Supplementary tables

**Supplementary Table 2:** Neutralisation in participants after the first dose of BNT162b2 vaccine against wild type and B.1.1.7, B.1.351 and P.1 spike mutant pseudotyped viruses.

|  | Number | Risk<br>ID50<20 | Unadjusted OR<br>(95% CI) | P value | Adjusted OR<br>(95% CI) | P value |
| --- | --- | --- | --- | --- | --- | --- |
| **WT** |  |  |  |  |  |  |
| **Age group years** |  |  |  |  |  |  |
| <80 | 55 | 25.5 (14/55) | 1 |  | 1 |  |
| ≥ 80 | 27 | 48.2 (13/27) | 2.7 (1.3-7.2) | 0.04 | 2.4 (0.8-6.8) | 0.06 |
| **Sex** |  |  |  |  |  |  |
| Male | 33 | 33.3 (11/33) | 1 |  | 1 |  |
| Female | 49 | 32.7 (16/49) | 1.0 (0.4-2.5) | 0.95 | 1.0 (0.3-3.2) | 0.94 |
| **Time since dose 1 weeks** |  |  |  |  |  |  |
| 3-8 | 28 | 25.0 (7/28) | 1 |  | 1 |  |
| 9-12 | 54 | 37.0 (20/54) | 1.8 (0.6-4.9) | 0.27 | 1.9 (0.6-6.0) | 0.25 |
| **Previous COVID-19** |  |  |  |  |  |  |
| No | 72 | 33.3 (24/72) | 1 |  | 1 |  |
| Yes | 6 | 50.0 (3/6) | 1.7 (0.4-10.7) | 0.42 | 1.8 (0.3-10.4) | 0.53 |
| **B.1.1351** |  |  |  |  |  |  |
| **Age group years** |  |  |  |  |  |  |
| <80 | 55 | 69.1 (38/55) | 1 |  | 1 |  |
| ≥ 80 | 27 | 81.5 (22/27) | 2.0 (0.6- 6.1) | 0.24 | 1.7 (0.5-5.7) | 0.41 |
| **Sex** |  |  |  |  |  |  |
| Male | 33 | 72.7 (24/33) | 1 |  | 1 |  |
| Female | 49 | 73.5 (36/49) | 1.0 (0.4-2.8) | 0.94 | 1.3 (0.4- 4.4) | 0.66 |
| **Time since dose 1 weeks** |  |  |  |  |  |  |
| 3-8 | 28 | 75.0 (21/28) | 1 |  | 1 |  |
| 9-12 | 54 | 72.2 (39/54) | 0.9 (0.3-2.5) | 0.79 | 1.0 (0.3-3.4) | 0.94 |
| **Previous COVID-19** |  |  |  |  |  |  |
| No | 72 | 77.8 (56/72) | 1 |  | 1 |  |
| Yes | 6 | 66.7 (4/6) | 0.6 (0.1- 3.4) | 0.54 | 0.5 (0.1-3.3) | 0.51 |
| **P.1** |  |  |  |  |  |  |
| **Age group years** |  |  |  |  |  |  |
| <80 | 55 | 52.7 (29/55) | 1 |  | 1 |  |
| ≥ 80 | 27 | 88.9 (24/27) | 7.2 (1.9-26.6) | 0.003 | 6.7 (1.7- 26.3) | 0.008 |
| **Sex** |  |  |  |  |  |  |
| Male | 33 | 66.7 (22/33) | 1 |  | 1 |  |
| Female | 49 | 63.3 (31/49) | 0.9 (0.3-2.2) | 0.75 | 1.2 (0.4-4.0) | 0.71 |
| **Time since dose 1 weeks** |  |  |  |  |  |  |
| 3-8 | 28 | 60.7 (17/28) | 1 |  | 1 |  |
| **9-12** | 54 | 66.7 (36/54) | 1.3 (0.5-3.3) | 0.59 | 1.5 (0.5-4.7) | 0.46 |
| **Previous COVID-19** |  |  |  |  |  |  |
| **No** | 72 | 68.1 (49/72) | 1 |  | 1 |  |
| **Yes** | 6 | 66.7 (4/6) | 0.9 (0.2-5.5) | 0.94 | 0.8 (0.1-5.5) | 0.77 |

**Extended data Figure 1:**
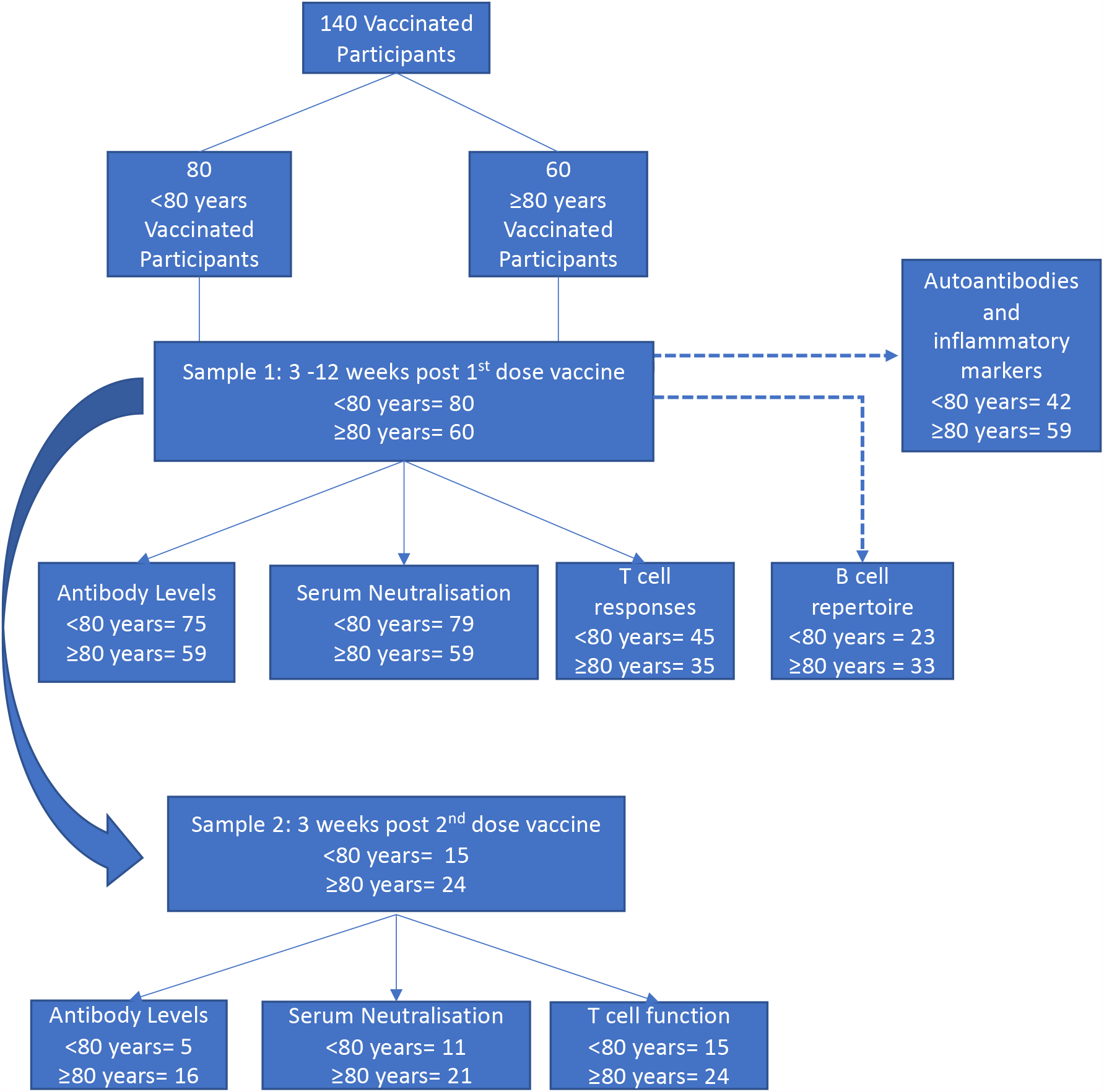
study flow diagram for samples and analyses

**Extended Data Figure 2.**
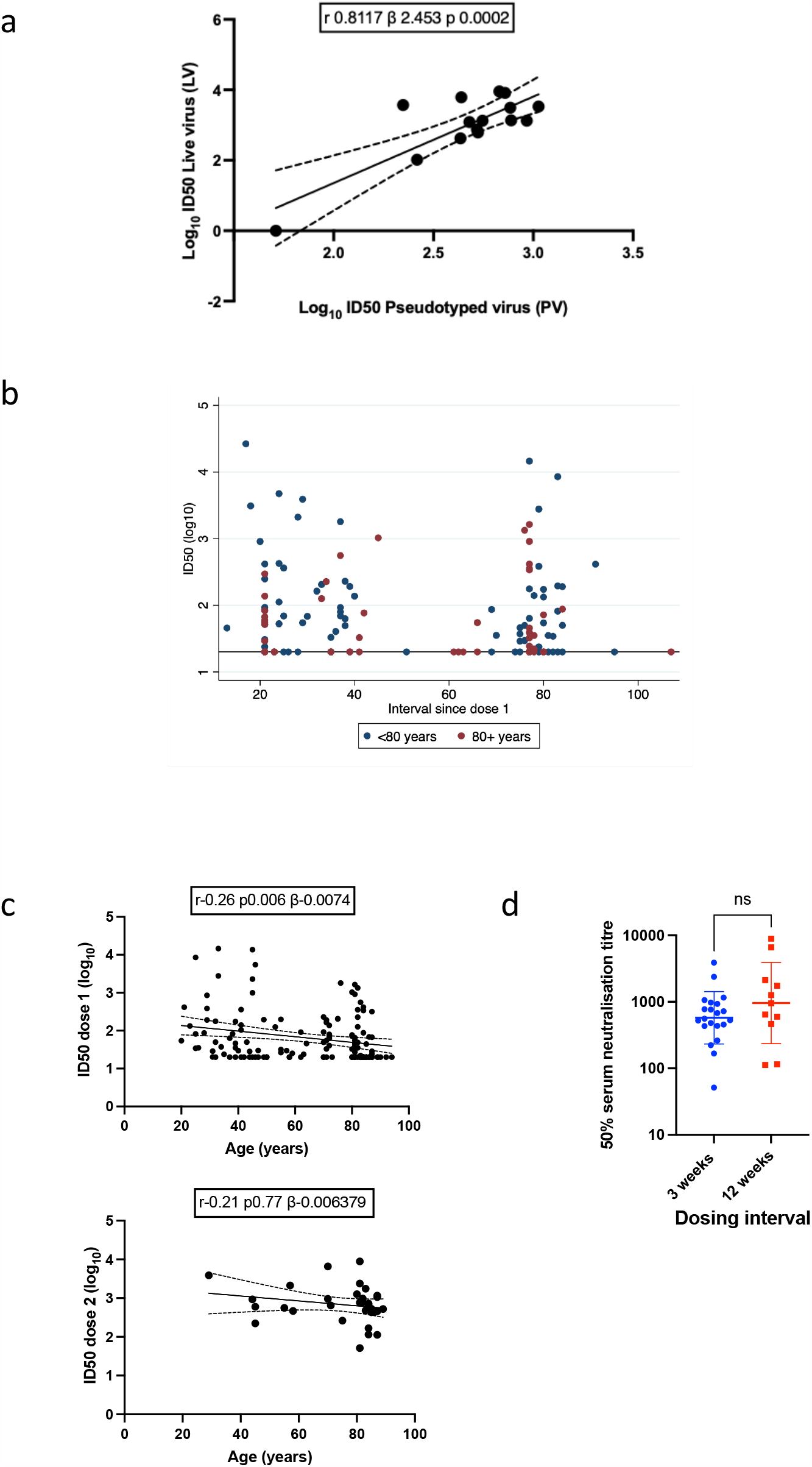
SARS-CoV-2 neutralisation by Pfizer BNT162b2 vaccine sera with age. **a**. Linear correlation of live virus neutralization with SARS-CoV-2 spike pseudotyped virus (PV) neutralization for 13 sera from individuals vaccinated with Pfizer vaccine. Linear regression line plotted bounded by 95% confidence interval. **b** SARS-CoV-2 PV neutralisation by Pfizer BNT162b2 vaccine sera following first dose in individuals (n=140) with time of sampling since dose shown on x axis. Red dots are individuals 80 years old and above, blue dots are those below 80 years old. **c**. Correlation of SARS-CoV-2 neutralisation by Pfizer BNT162b2 vaccine sera with age. Serum neutralisation of Spike (D614G) pseudotyped lentiviral particles (inhibitory dilution at which 50% inhibition of infection is achieved, ID50) after Dose 1 (A n=138) or dose 2 (B n=32) by age. Linear regression line plotted bounded by 95% confidence interval. r– Pearson’s correlation coefficient, β slope/regression coefficient, p p-value. Bonferroni adjustment was made for multiple comparisons n linear regression. **D**. ID50 against WT (D614G) pseudotyped virus (PV) following the second dose of vaccine stratified by interval between vaccine doses [3 weeks (n=21) and 12 weeks (n=11)]. Geometric mean titre with s.d. Mann-whitney test.

**Extended Data Figure 3.**
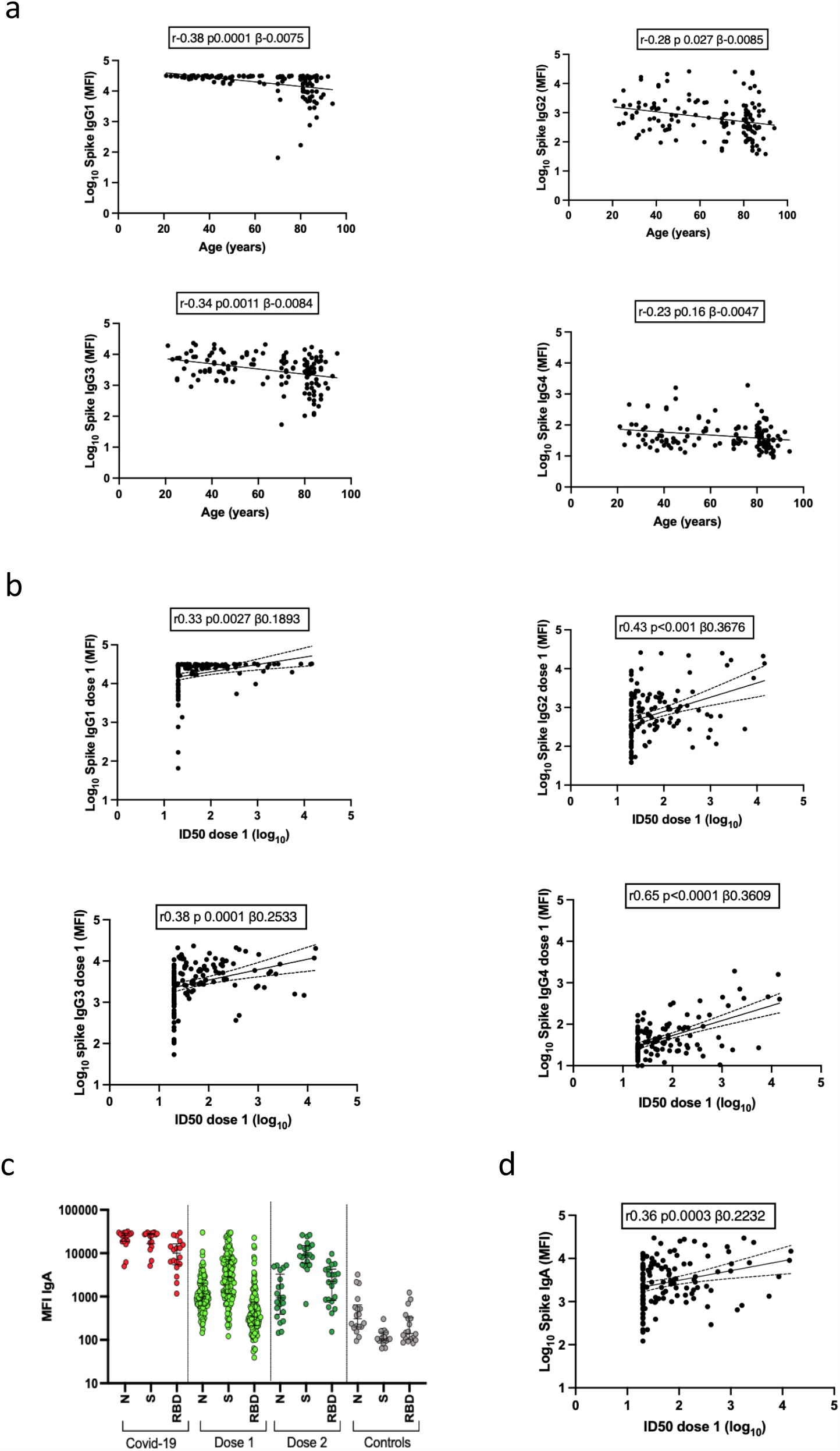
Binding IgG and IgA spike antibody responses following BNT162b2 vaccination. **a**. Correlations between serum binding IgG subclass 1-4 antibody responses following vaccination with first dose of Pfizer BNT162b2 vaccine and age in years (n=133). **b**. Correlations between serum binding IgG subclass 1-4 antibody responses following vaccination with first dose of Pfizer BNT162b2 vaccine and serum neutralization using a pseudotyped viral (PV) system (n=133). **c**. IgA responses to S, N, RBD post first dose (light green, n=133) and second dose (dark green, n=21) compared to individuals with prior infection (red, n=18) and negative controls (grey, n=18) at serum dilutions 1 in 100.**d**. Correlations between serum binding IgA spike antibody responses following vaccination with first dose of Pfizer BNT162b2 vaccine and age in years and serum neutralization using a pseudotyped viral system (n=133). MFI-mean fluorescence intensity ID50 – inhibitory dilution required to achieve 50% inhibition of viral infection. r pearson’s correlation coefficient, β slope/regression coefficient, p p-value. Bonferroni adjustment was made for multiple comparisons. Spike proteins tested are Wuhan-1 with D614G.

**Extended Data Figure 4:**
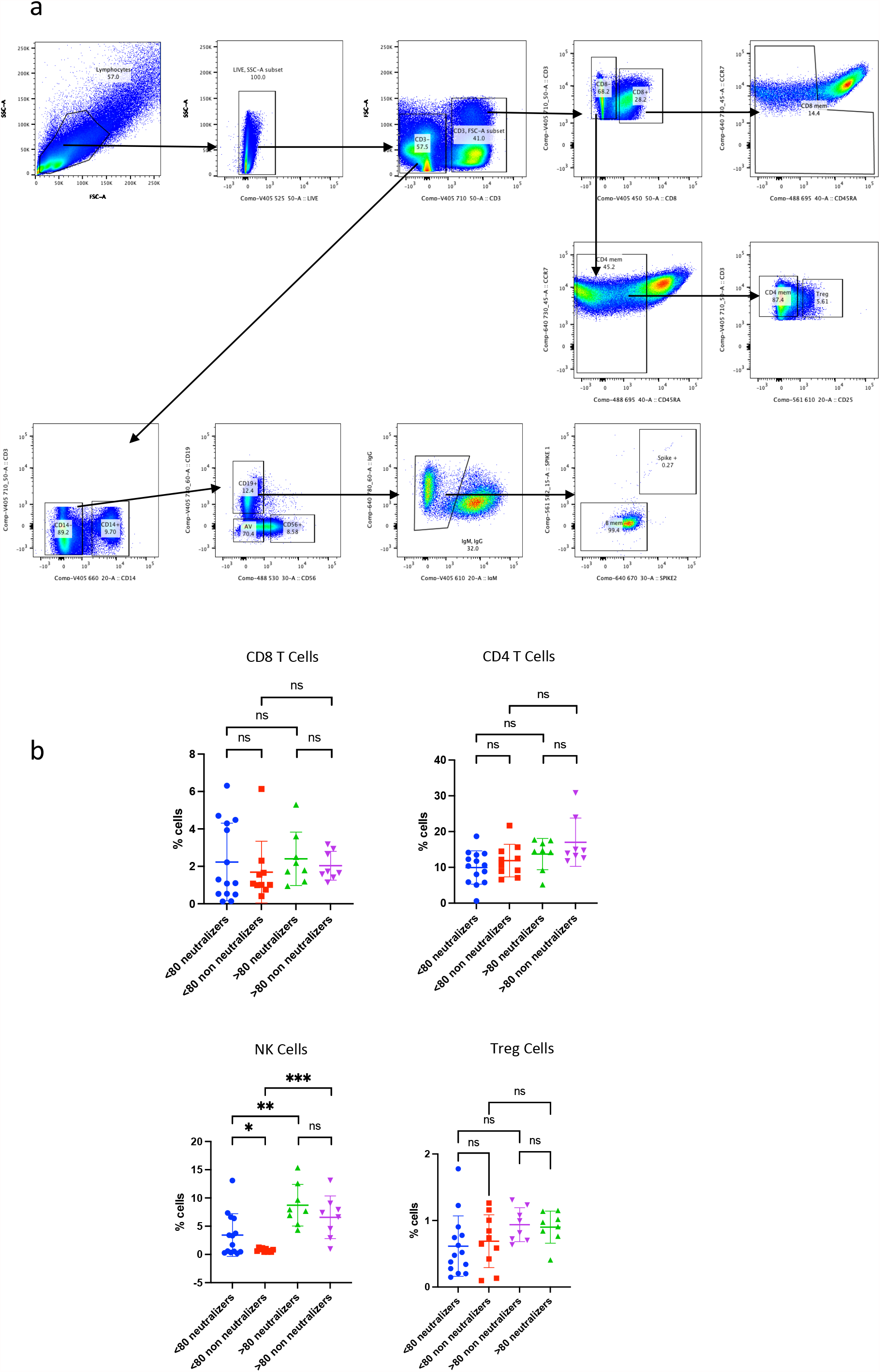
Peripheral blood Lymphocyte subsets following first dose BNT162b2 vaccination. PBMC were FACS sorted (n=16 above 80 and n=16 below 80). **a**. Gating strategy for flow cytometry analysis of human immune cells post BNT162b vaccination. **b** data for indicated sorted cell subsets stratified by neutralizing response after first dose, n=8 in each category). NK cell CD3- CD14- CD19 - and CD56 positive cells

**Extended Data Figure 5.**
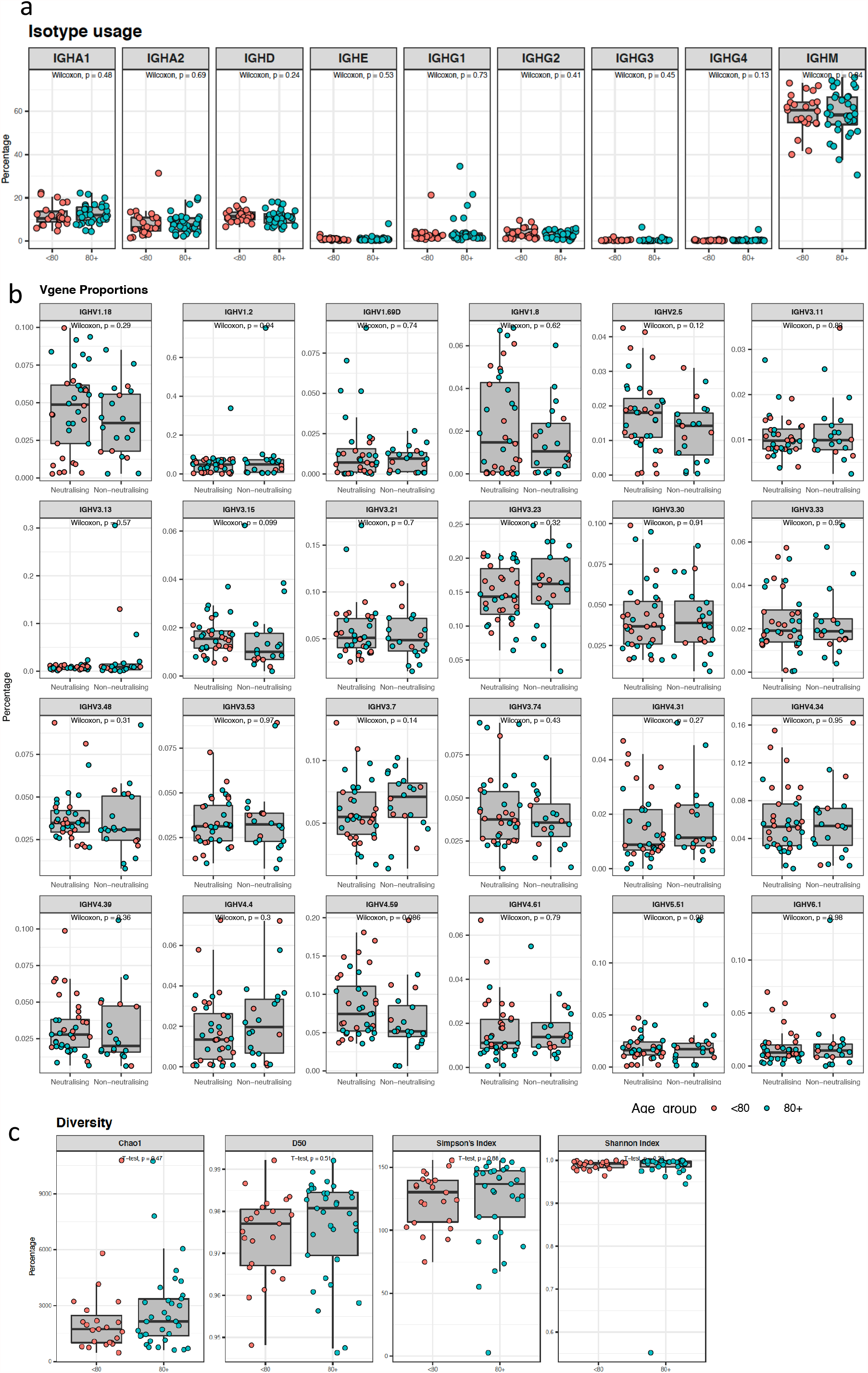
B cell repertoire following vaccination with first dose of Pfizer BNT162b2 vaccine. **a**. Isotype usage according to unique VDJ sequence comparing under 80 year olds with 80 year olds and older. **b**. Boxplots showing V gene usage as a proportion, comparing neutralisation of spike pseudotyped virus. Neutralisation cut-off for 50% neutralisation was set at 20. **c**. Diversity Indices comparing under 80 year olds with 80 year olds and older. The inverse is depicted for the Simpson’s index and the Shannon-Weiner index is normalised.

**Extended Data Figure 6.**
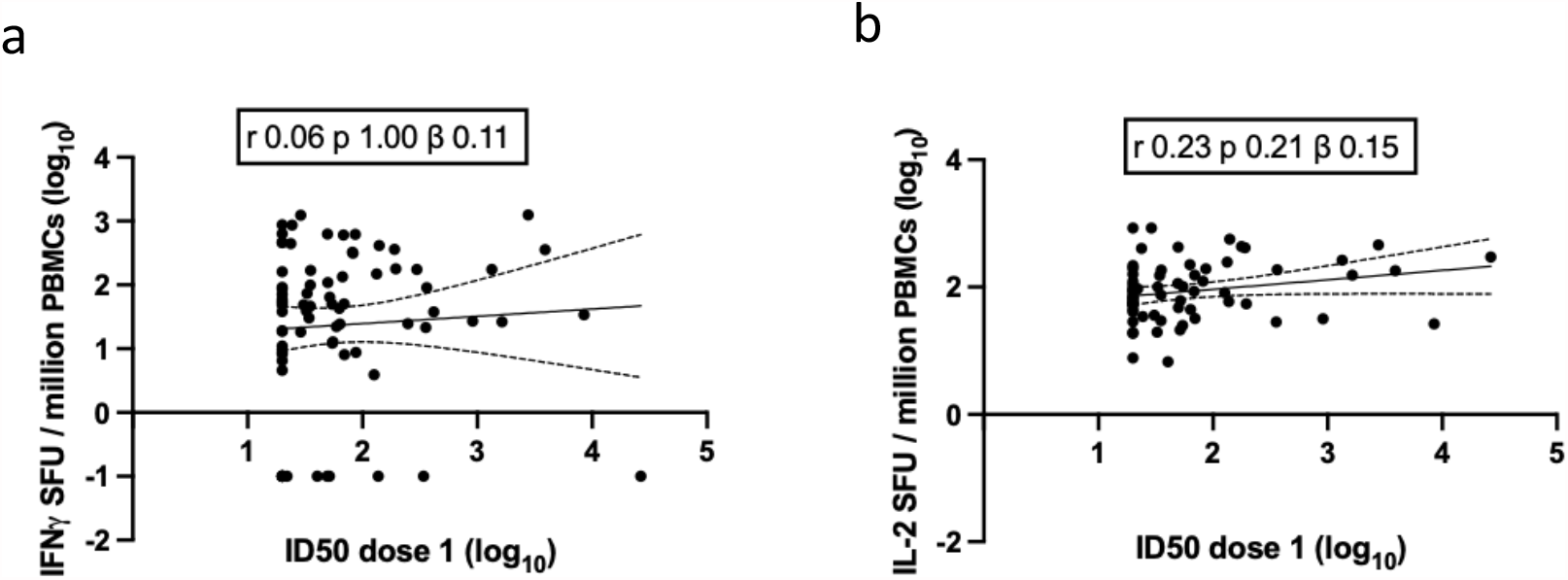
Correlation between T cell responses against SARS-CoV-2 Spike peptide pool and serum neutralisation of Spike (D614G) pseudotyped lentiviral particles (inhibitory dilution at which 50% inhibition of infection is achieved, ID50). **a**,**b**. Correlation of IFNγ (n=79) and IL2 (n=69) FluoroSpot and ID50 after first dose. SFU: spot forming units. Linear regression line with 95% confidence intervals are plotted. r: Pearson’s correlation coefficient. p value indicated and b the slope or coefficient. Bonferroni adjustment was made for multiple comparisons.

**Extended Data Figure 7.**
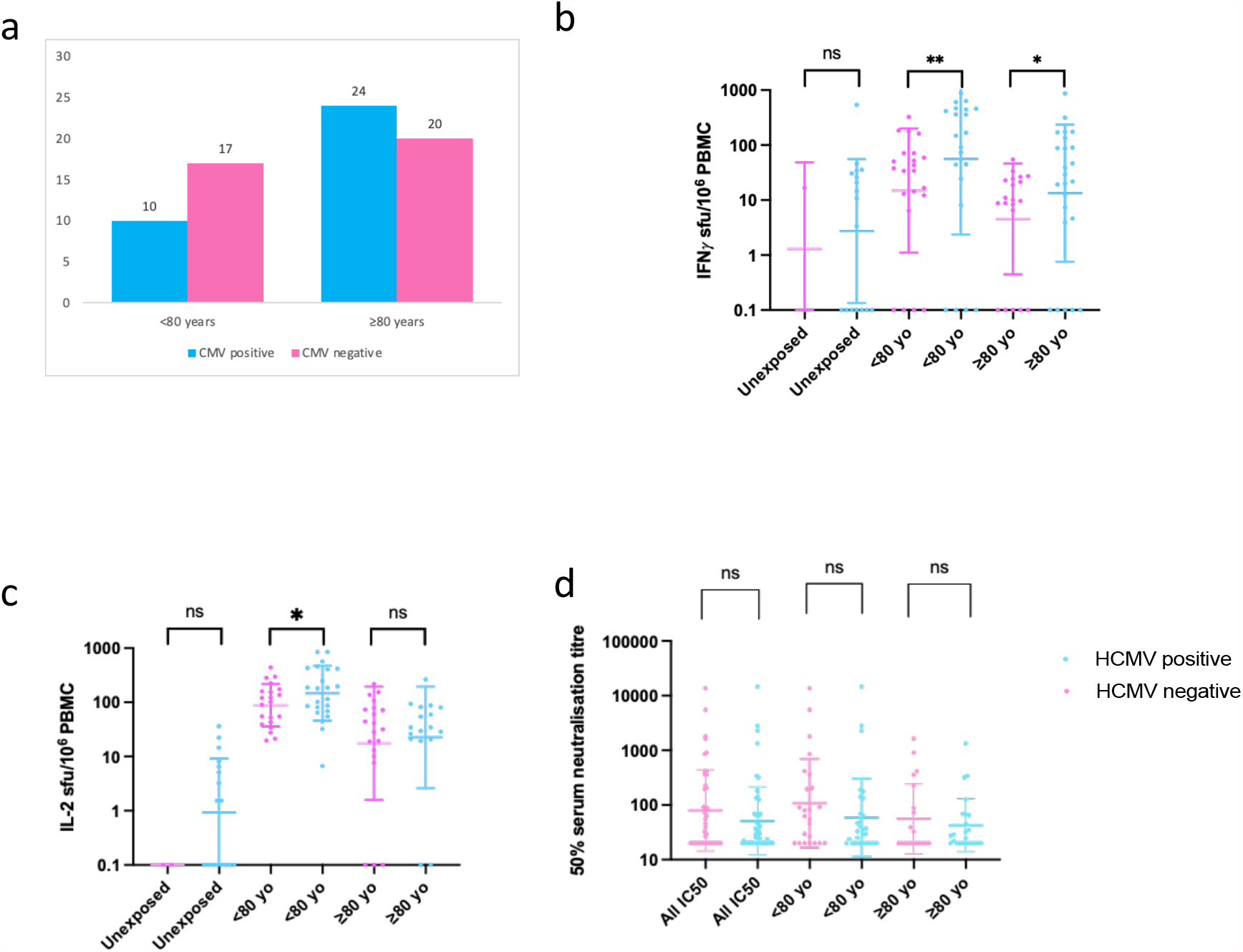
Human cytomegalovirus serostatus, T cell responses and serum neutralisation of Spike (D614G) pseudotyped lentiviral particles (inhibitory dilution at which 50% inhibition of infection is achieved, ID50) to Pfizer BNT162b2 vaccine after the first dose of vaccine. Dose 1. **a**. (n=72) HCMV serostatus by <80 and ≥80 year age groups, HCMV positive (blue), HCMV negative (pink). **b** IFNγ (n=72) and **c**. IL2 (n=64) FluoroSpot response after the first dose. **d**. Inhibitory dilution at which 50% inhibition of infection after the first dose. SFU-spot forming units. ID50-inhibitory dilution at which 50% inhibition of infection is achieved. HCMC-Human cytomegalovirus., CEF-Cytomegalovirus Epstein Barr virus, Influenza virus

**Extended Data Figure 8:**
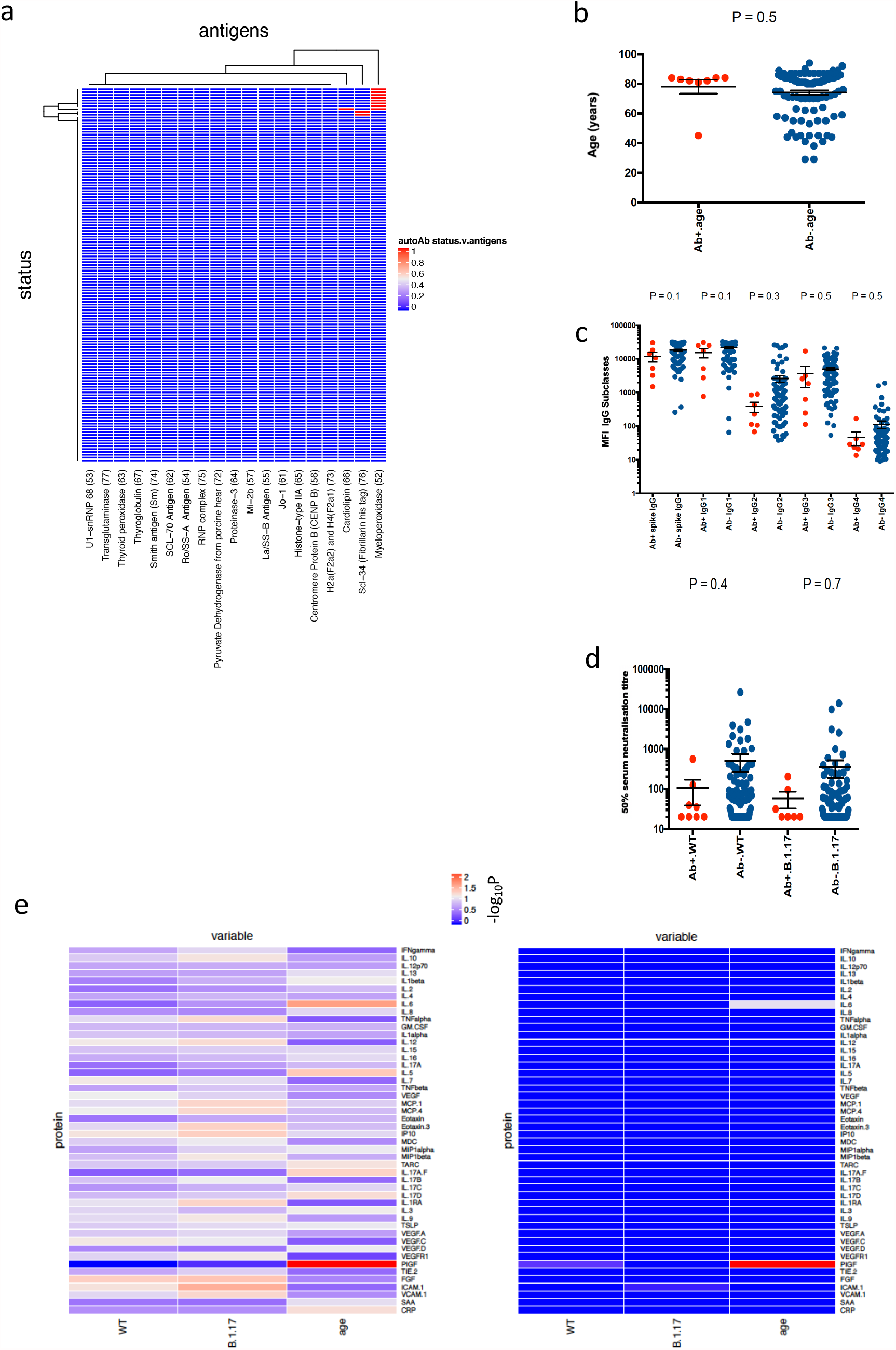
Autoantibodies and inflammatory markers in participants receiving at least one dose of the Pfizer BNT162b2 vaccine and relationship to SARS-CoV-2 spike specific IgG and SARS-CoV-2 PV neutralisation. (n=101). **a**. Heatmap of log2 transformed fluorescence intensity(FI) of 19 autoantibodies, positive (red), negative (blue). **b**. Age in years by anti-MPO antibody positive (red) or negative (blue) status. Plotted is the mean age and s.d. **c**. (n=100) IgG subclass responses to Spike post first dose Pfizer BNT162b2 vaccine comparing individuals with anti-MPO antibody positive (red) or negative (blue) status. **d**. GMT with s.d of first dose Pfizer BNT162b2 vaccine sera against wild type and B.1.1.7 Spike mutant SARS-CoV-2 pseudotyped viruses by anti-MPO antibody positive (red) or negative (blue) status. Ab+ antibody positive, Ab-antibody negative, MPO-myeloperoxidase, P – pvalue, WT-wild type, B.1.1.7 Spike mutant with N501Y, A570D, ΔH69/V70, Δ144/145, P681H, T716I, S982A and D1118H. **e**.Nonparametric rank correlation (Kendall’s tau-b) of wild type (WT), variant (B.1.17) and age (<>80 years) against each of 53 cyto/chemokines. Heatmaps illustrate Tau-b statistic (left) and significance (right, –log_10_FDR).

